# Seasonality, source type, and women’s water labor: A longitudinal mixed-methods study in Kenya and Honduras

**DOI:** 10.64898/2026.06.09.26355008

**Authors:** Thea Mink, Emily Ogutu, Madeleine Patrick, Sheela S. Sinharoy, Margot V. Bolaños Gamez, Alicia Macler, Christopher P. Ngo, Harriet Oglesby, Olivia Bendit, Jera White, Sandra Antonio, Gladys Ramos, Edwin Roldan Medina Lopez, Everlyne Atandi, Peter Mwangi, Peter Koome, Rohin Otieno Onyango, Petronilla Andiba Otuya, Paul Ruto, Bethany A. Caruso

## Abstract

Women shoulder the majority of water collection labor globally, yet how their water collection and water-related work experiences may change over time or by water source type remains insufficiently understood. We conducted a longitudinal, mixed-methods study in rural Kenya and Honduras to understand how women’s experiences collecting water and performing water-related work varied between (a) two time points, (b) improved and unimproved water source types, and (c) water source location.

Data were collected in 2023 and 2024 using interviews, observation, GPS-enabled watches, and scales to measure time and distance traveled, water weight and volume carried, and calories expended. 133 women participated in data collection (66 Kenya, 67 Honduras). We compared women’s experience data by time point (2023 vs. 2024), source type (improved vs. unimproved), and source location (off-premises vs. on-premises) (t-test, Mann-Whitney U test). We also mapped participants’ routes and activities to show which sources were visited, when, and for what activities.

In Kenya, mean water collection time, distance, and caloric expenditure were significantly lower and water volume was significantly higher in 2024 when there were unexpected rains compared to 2023 when there was a persistent drought. When comparing source types during the 2023 drought, journeys to improved sources took significantly less time and energy and covered less distance than journeys to unimproved sources. These differences were not observed during the rainy conditions of 2024 when unimproved sources were closer and more accessible. In Honduras, water collection and water work burdens did not differ significantly by time point or source type. We found women with on-premises water access to still expend considerable time and caloric expenditure engaging in water work within their household compounds.

Findings from Kenya suggest that water infrastructure improvements can reduce women’s water collection burdens, though benefits may depend on and vary by season and source location. Findings from Honduras show that water labor does not end once water is in the household. Rather, substantial time and energy are expended carrying out water-related work even when sources are on premises, suggesting that efforts to assess water labor need to extend beyond collection alone. To meaningfully reduce burdens and ensure improved water sources are utilized during all seasons, initiatives need to consider source location, seasonal variability, and work beyond collection. Evaluations to assess infrastructure impacts on women’s labor and well-being are needed and long overdue.

## 1 INTRODUCTION

While global coverage of drinking water is increasing, an estimated 1.7 billion people still do not have access to water within their household premises [1]. In many countries, the tasks of collecting and carrying water continue to be gendered; globally 70% of this burden is shouldered by women (63%) and girls (7%) [1, 2]. It is estimated that women and girls spend 250 million hours collecting water each day, over three times the time spent by men and boys [3]. In addition to time, the demands of water collection and water-related work for women include costs to their energy, physical and mental health, and opportunities, such as economic engagement, leisure time, and sleep [4–12]. However, existing research has been largely cross-sectional, limiting understanding of how experiences vary over time and across source types and locations. Strengthened understanding of how women’s water collection and water labor experiences may vary over time or change as water source access improvements are implemented is therefore essential for assessing progress in both water service delivery and women’s health and well-being.

Improved water access is foundational for direct health benefits and can contribute to broader outcomes for women’s lives. Improved water sources like piped water, boreholes, protected dug wells, and protected springs have the potential to deliver safe water [13] and water from these sources has been associated with a reduced risk of diarrheal diseases [14], improved child linear growth [15], and lower maternal mortality [16, 17]. With improved access to water, women have reported less psychosocial stress, greater well-being, and improved household relationships [18–20]. Other benefits of water access can include time savings, which may be reallocated to rest, leisure, other household work, or income-generating activities [18, 21–23]. A systematic review and meta-analysis of water collection travel time found that mean time decreased 15 minutes per trip for water supply interventions, or about eight hours per week, with the greatest mean reductions in sub-Saharan Africa (21 minutes per trip) [21]. In an analysis of 26 countries in Africa, a 15-minute decrease in roundtrip water fetching was associated with reductions in child morbidity, potentially also reducing the time women dedicate to caring for sick children [24]. These findings highlight the multidimensional benefits of improved water access, particularly for women.

However, when water sources are improved but remain off household premises, positive impacts on women’s daily lives are not assured. While placing water sources closer to homes has the potential to reduce travel distance and time per trip, women may still encounter new or intensified demands, including carrying larger volumes, queuing for long periods, making more frequent trips, paying fees at sources, or adjusting their schedules to source operating times [23, 25–27]. For example, a qualitative study in northwest India found that installing public piped water reduced the distance women walked but increased the volume of water they were expected to retrieve for men’s and household hygiene [25]. Similar patterns have been observed in Benin and Indonesia, where improved water access led to more frequent trips, time waiting in line, and expectations that women fetch [23, 26]. In rural Zambia, new borehole installation reduced women’s time for collection, however the responsibility was displaced onto girls; because the source was closer, mothers felt it was now safer for girls to collect water [27]. As a result, while burden decreased for women, girls’ water collection time significantly increased and their time at school decreased [27]. Moreover, having access to an improved source does not guarantee its use. Women may decide to use other unimproved sources if an improved source is unavailable [4], requires a fee [23], or does not meet their specific needs (e.g., have a flow rate suitable for washing) or preferences (e.g., provides water that is palatable for drinking) [28, 29]. In addition to not guaranteeing positive impacts on women, this existing research shows that providing improved water infrastructure may even increase women’s water collection burdens.

Further research is needed to assess women’s experiences collecting water, including after water improvements are introduced and across different seasons when water source availability may fluctuate. To conduct this work, there is a need for approaches that accurately and reliably capture quantitative changes in women’s water collection experiences, including time, distance, and energy. Time data are typically collected through self-report (e.g., national household surveys), which is appropriate for national and global monitoring [30–32], yet self-reported time has been shown to both overestimate and underestimate actual collection time when compared to observed time [4, 33–36]. Although time is often used as a proxy for distance, it is an imprecise stand-in because factors like elevation, terrain, physical ability, load weight, and activities at or along the way to a source can influence trip duration and distance estimates [4, 35, 37, 38]. The few studies that have used Geographic Information System (GIS) to measure straight-line, Euclidean distance from home to water sources suggest that the method better estimates distance than self-reported time but is also less accurate than Global Positioning System (GPS) measures of distance [35, 37, 39]. Further, data on women’s energy expenditure for water collection is limited [4, 40, 41].

In an earlier study, we sought to improve upon previous assessments of women’s experiences by using go-along interviews, GPS-enabled watches, and scales to document time and distance traveled, energy expended, weights and volumes carried, and activities performed by women collecting water outside the household in rural Guatemala, Honduras, Kenya, and Zimbabwe [4]. Although this previous work provided a rigorous and reliable approach for capturing women’s experiences, data were only collected in a narrow window of time in each country setting. As a result, we could not determine if women’s experiences would be the same if data were assessed at another time.

The present study returned to two countries where research had previously been conducted, Honduras and Kenya, to further understanding of women’s water collection experiences. We collected new data in 2024 and leveraged previously collected data from 2023 to understand if there were differences in water collection experiences at each time point and by type of water source visited (improved vs. unimproved). We aimed to answer the following three research questions:

a. Are women’s water collection experiences different in 2023 compared to 2024?
b. Are women’s water collection experiences different among those who collect water from improved sources compared to those who collect water from unimproved sources?
c. How does women’s water-related work differ between those who use water from on-premises sources compared to those who collect water from off-premises sources?

## 2 METHODS

### 2.1 Study design

This longitudinal, observational, mixed-methods study used (a) go-along, in-depth interviews (IDIs), (b) semi-structured observation, (c) GPS-enabled watches, and (d) weighing scales to understand women’s experiences going to water sources (**Fig 1**) [4]. Research was conducted in Honduras and Kenya with communities participating in the World Vision program, Strong Women Strong World: Beyond Access (SWSW: BA), in 2023 and 2024.

**Fig 1.**
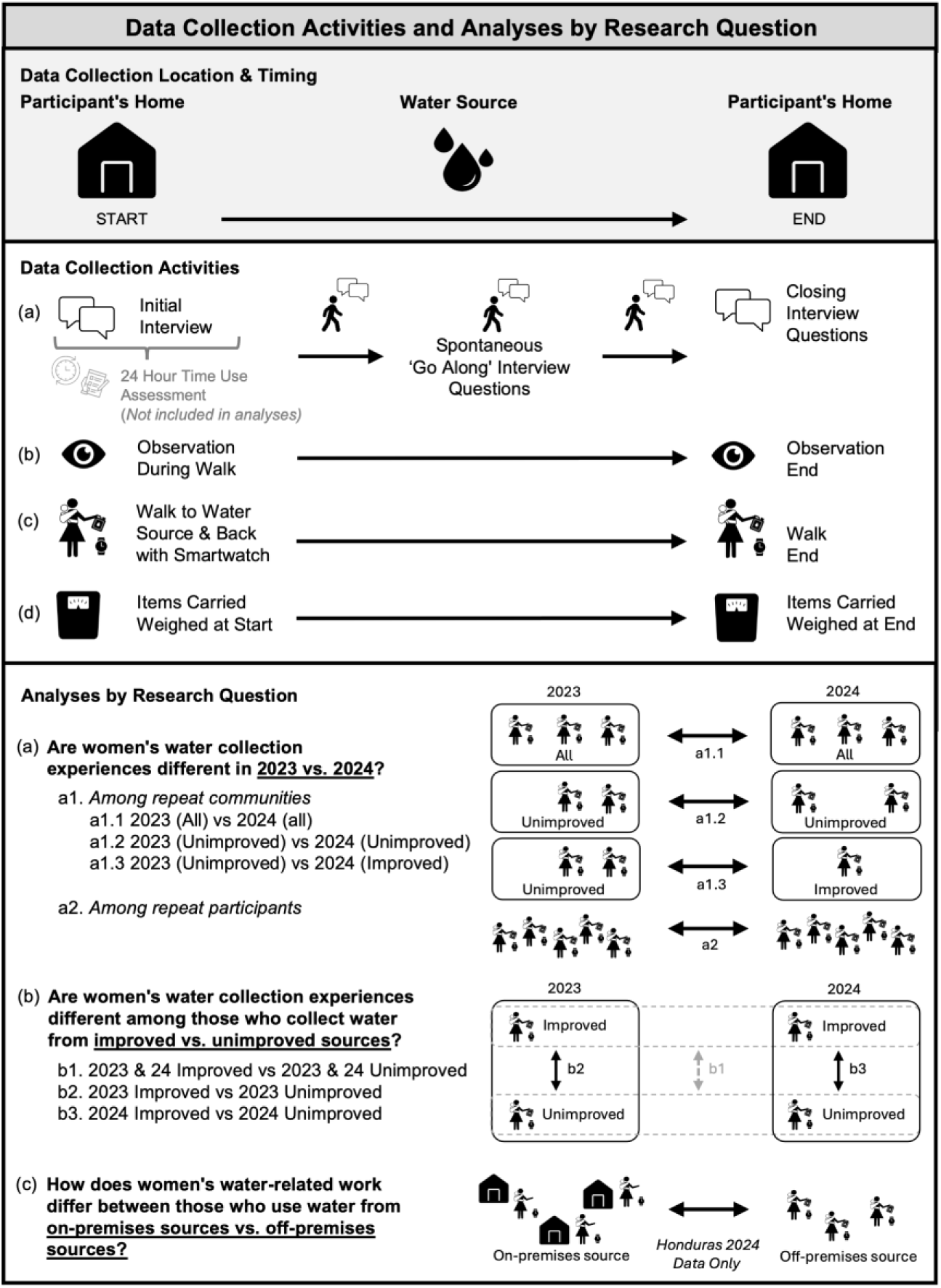
Data collection activities and analyses by research question. Adapted from: Women’s experiences collecting and accessing water in Guatemala, Honduras, Kenya, and Zimbabwe: A mixed-methods investigation. *PLOS Glob Public Health* 5(12): e0004355, CC BY 4.0.\

The SWSW: BA program aims to strengthen women’s empowerment through an integrated delivery of improved water resources and a suite of economic empowerment activities. The program theorizes that improved water access will save women time and energy and improve water availability, thereby allowing them to redirect these resources toward economic activities. Emory University partnered with World Vision US and World Vision SWSW: BA teams in Honduras and Kenya to design this study. The World Vision Honduras and Kenya teams identified local research partners in each country that led data collection: Universidad Nacional Autónoma de Honduras (UNAH, 2023 partner) and the Edwin Roldan Medina Lopez Research Group (2024 partner) in Honduras and St. Paul’s University in Kenya (2023 and 2024 partner).

### 2.2 Setting and sample

Communities were purposively selected in partnership with World Vision Kenya and World Vision Honduras teams based on their participation in the SWSW: BA program.

Kenya data collection occurred in ten rural communities located in Samburu and Isiolo counties. Data collection occurred at similar time points each year (June-July 2023, July-August 2024). The region is arid and semi-arid and regularly experiences drought. During 2023 data collection, the area was experiencing a prolonged drought [42]. While data collection in 2024 was also anticipated to occur during the dry season, the region experienced heavy rainfall earlier than expected. Of the total population in Kenya, 66% have at least basic water service (56% rural, 89% urban) [2]. Honduras data collection occurred in nine rural and peri-urban communities located in Danli and Teupasenti municipalities. Both regions are forested and have a tropical climate. Data collection occurred during the rainy season at both time points (July-August 2023 and 2024). Of the total country population, 96% have at least basic water service (90% rural, > 99% urban) [2].

For the first year of data collection in 2023, we included six communities in each country setting to reflect geographic and population diversity (**Table 1**). In Kenya, three communities had access to an improved water source (borehole) that was installed in one of the communities by World Vision before data collection. In Honduras, none of the communities had received improved water sources from World Vision before data collection.

**Table 1.** Community and participant engagement for Kenya and Honduras data collection in 2023 and 2024.

|  | Kenya |  | Honduras |  |
| --- | --- | --- | --- | --- |
|  | 2023 | 2024 | 2023 | 2024 |
| **Communities engaged in data collection** |  |  |  |  |
| Total (new, repeat)<sup>1</sup> | 6 | 6 (4 new, 2 repeat) | 6 | 6 (3 new, 3 repeat) |
| Improved water delivered<sup>2</sup> | 3 | 3 (1 new, 2 repeat) | 0 | 2 (0 new, 2 repeat) |
| No improved water delivered<sup>3</sup> | 3 | 3 (3 new, 0 repeat) | 6 | 4 (3 new, 1 repeat) |
| **Participants engaged in data collection** |  |  |  |  |
| Total<sup>4</sup> | 22 | 48 | 17 | 51 |

<sup>1</sup> New communities were included for one year of data collection; repeat communities were included for data collection in both 2023 and 2024.
<sup>2</sup> Improved water source was installed by World Vision before data collection.
<sup>3</sup> Improved water source was not installed by World Vision before data collection. Some participants had access to other improved water sources.
<sup>4</sup> Total includes new and repeat participants. 133 unique participants were engaged in data collection; 5 were repeat participants (4 Kenya, 1 Honduras).

For data collection in 2024, we again included six communities in each country setting (**Table 1**). We included ‘repeat communities’ that were included in 2023 data collection, as well as ‘new communities’ that were not included in earlier data collection. Repeat communities that received improved water resources from World Vision had infrastructure installed after 2023 data collection, but before 2024 data collection. In Kenya, two repeat communities and one new community received improved water infrastructure (public water kiosk) from World Vision before 2024 data collection. In Honduras, two repeat communities received improved water infrastructure (household piped water) from World Vision before 2024 data collection, and a third repeat community was in the process of receiving improved water infrastructure from World Vision during 2024 data collection.

For both years of data collection, participants were eligible to participate if they were women, aged 18 years or older, responsible for collecting water for household needs, and lived in eligible communities. Both years, we used purposive and convenience participant sampling. For participant selection in 2023, we aimed to include at least three women per community for a total minimum sample of 18 participants for each country setting; a total of 39 were included across all country sites, including 22 in Kenya and 17 in Honduras (**Table 1**). For participant selection in 2024, we aimed to recruit and enroll at least eight women per community for a total minimum sample of 48 participants per country setting to increase variability; 99 participants were engaged (48 in Kenya and 51 in Honduras) including five repeat participants (**Table 1**). Repeat participants were identified in 2024 using contact information and demographic survey data provided by 2023 participants who expressed interest in being recontacted for follow-up research.

Recruitment varied slightly between Honduras and Kenya both years. In Kenya, the local research team met with village leaders to explain the research project and ask for their help in identifying possible participants. The leaders then spoke with community members to gauge interest and relayed this information to the research team. The research team followed up by visiting those who expressed interest to further explain the study and consent interested and eligible participants. World Vision Kenya staff did not participate in recruitment.

For recruitment in Honduras, the local World Vision team connected community leaders (e.g., members of the boards of trustees, water boards, churches) with the local research team. The local research team explained the study to the leaders, who then guided them through the community. The local research team went door to door to share information about the study and invited eligible and interested women to participate. During recruitment, women were not asked if they had access to improved water sources at home or off premises, so as to not influence their water source selection during data collection. World Vision Honduras staff did not participate in recruitment beyond introducing local learning partners to community leaders.

### 2.3 Data collection procedures and tools

Data collection occurred in 2023 (Honduras: July 5-August 5; Kenya: June 22-July 24) and 2024 (Honduras: July 22-August 8; Kenya: July 10-August 14). In both study areas and years of data collection, women chose where to go for their water source.

#### 2.3.1 Qualitative data collection

To understand women’s own perspectives and experiences about water collection and water work at the sources, go-along in-depth interviews were carried out (**Fig 1**). Go-along interviews enable participants and interviewers to discuss the environment they are walking through and encourage participants to take an active role by guiding the interviewer and reflecting on the space [43, 44].

Women were also explicitly asked what water source they used during data collection, how many total times they anticipated using the source that day, and the time they anticipated needing to manage water once they collected it (see full guide in **S1 Tools**).

To further document the experiences and conditions within which women were venturing to water sources, data collectors conducted semi-structured observations (see full guide in **S1 Tools**). Observation guides prompted data collectors to record information about, for example, the water sources visited, terrain traversed, how women extracted and transported water, and activities women carried out while going to, collecting water at, and returning from the sources. In Honduras, photos were taken of the water sources used during data collection to help identify the source type.

#### 2.3.2 Quantitative data collection

To record total time, distance, and caloric expenditure, participants wore Garmin Forerunner 255 watches (**Fig 1**). Watches were calibrated using each participant’s weight, height, and age to estimate caloric expenditure; caloric expenditure is reported in kcal. A data collection team member also wore a watch as a back-up recording, and their watch was calibrated using the participant’s weight, height, and age. The participants’ and data collectors’ watches were clearly distinguishable and not interchanged to maintain data separation. Data collectors started the watch recording when the participant left their home and ended when they returned. However, in Honduras during the 2024 data collection, many women remained within their household compound or yard because they had piped water access (as mentioned above, women were not asked at screening about where they would get their water that day so as to not bias their behaviors). Therefore, data collectors started watches when participants started water-related tasks (e.g., washing clothes, cleaning water storage containers) and ended when the tasks were completed.

To determine the weight of carried items, Seca 876 scales were used (**Fig 1**). Data collectors weighed items that each woman carried at the start of her trip, such as empty water containers, a child, or dry laundry. At the water source, items were re-weighed before participants started their return trip, such as water containers with water or wet laundry. Any other items that were picked up along the way (e.g., firewood) were also weighed. If items were difficult to place on the scale alone (e.g., laundry), data collectors weighted the woman while holding the item(s) and later subtracted her weight. For Honduras 2024 data collection, when many women remained on premises for data collection, water was weighed if women retrieved it from *pila* storage containers for use.

To gain basic information about each participant (e.g., demographic characteristics, their primary source for drinking water), participants responded to a brief survey that was completed at the participant’s home before other activities began. The survey tool can be found in **S1 Tools**.

### 2.4 Data collection training

Emory University team members trained Honduras and Kenya-based research teams on the data collection tools and methods for this study. The training topics included information about the SWSW: BA program goals, the conduct of ethical research, qualitative and quantitative research principles, data management, and how to use the research tools and hardware (e.g., watches, scales, and recorders).

Tool translations were reviewed by the local research teams for consistent meaning in English, Samburu, and Spanish. Before data collection began, all tools and data collection procedures were piloted in communities similar to those to be visited for the study (Kenya) or within the enumerator teams (Honduras).

### 2.5 Data collection and management

Data were collected by pairs of trained women research team members. One data collector conducted the survey with the participant, explained and set up the activity tracking watch, and facilitated the go-along in-depth interview. The other team member recorded notes for the semi-structured observation and weighed all items that the participant carried. Data collection was conducted in Samburu (Kenya) and Spanish (Honduras).

Digital audio recorders were used to record the go-along in-depth interviews. In Kenya, recordings were simultaneously translated and transcribed from Samburu into English and validated by the trained research team. In Honduras, audio recordings were transcribed into Spanish by the local research team and then translated to English by a hired translator in 2023 and by a trained translation team in 2024. To ensure data quality, translated transcripts were cross-checked by team members who were not involved in the initial transcription or translation.

Survey data and observation notes were recorded on paper. In Honduras, interview and observation notes were recorded in Spanish, which were then translated into English by a hired translator. In Kenya, interview and observation notes were recorded in English. To ensure data accuracy, survey and weight data were reviewed by the local learning partners, double entered independently by two members of the Emory team, and then compared for accuracy. Any discrepancies were checked against the scanned forms and corrected.

Watch data were synced to the ‘Garmin Connect’ platform and named with participant IDs. Data were downloaded from the platform as CSV and GPX files and then uploaded to a secure folder on OneDrive. To confirm that the data were for the correct participant and that the recordings were complete, participant watch data (e.g., start and end times) were compared to that of the data collector. Data were deleted from the watches after being uploaded.

### 2.6 Analysis

To determine actual water collection and water-related work time, distance, caloric expenditure, water volume, and water volume per household member, we calculated descriptive statistics of watch and weight data. Water volume was determined from the weight of water alone (1 kg = 1 L), which was calculated by subtracting the weight of empty water containers from the weight of the containers with water. Water volume per household member was calculated by dividing water volume by the number of household members participants reported in the survey. The weight of water for seven participants (three in Kenya and four in Honduras) included the weight of container, because the weights of the participants’ containers without water were not recorded. (From previous work, mean empty water container weight was 1.4 kg in Kenya and 0.7 kg in Honduras [4].) Estimated water labor time for the day of data collection was calculated by multiplying the watch-measured time of a participant’s journey to the water source and back by the number of trips the participant reported they planned to make to that water source on the day of data collection. The estimated water labor time for the day was only calculated for 2024 data as participants were not asked about the number of times they anticipated going to the source in 2023.

Time, distance, and caloric expenditure were manipulated for one participant in Kenya and one participant in Honduras because of data collection challenges: watch data for the data collectors were used because the participants’ watches did not record. Additionally, one Honduras participant’s metrics were doubled as they were accidentally only recorded for one-way.

To determine what type of water source participants used during data collection, we used observation notes and go-along interview transcripts that described the source. Water source types were categorized using the WHO/UNICEF Joint Monitoring Programme for Water Supply, Sanitation, and Hygiene (JMP) definitions for unimproved and improved sources [45]. In Honduras, *pilas,* or open water containers often filled with piped water, were common within household compounds. We categorized participants as using piped water if they used their household *pila* and reported in the survey that they used piped water as their primary source for drinking water. When possible, we confirmed source types by reviewing photos taken of the water sources. Additionally, we determined what activities were carried out when women were going to and from water sources using observation notes and go-along interview transcripts.

To determine participants’ estimated total number of trips to the water source for the day and participants’ estimated time to manage water at home for the day, we referred to the interview transcripts. These data were only collected in 2024.

We used the following analytical approaches to compare data on women’s experiences in alignment with our research questions (**Fig 1**).

#### 2.6.1 Research Question a: Are women’s water collection experiences different in 2023 compared to 2024?

To answer this question, we analyzed data from two groups of participants. One group consisted of participants who lived in communities that were engaged in both 2023 and 2024 (repeat communities). The other group included participants who participated in both years (repeat participants).

##### Repeat community analysis

The analytic sample for this analysis included participants in communities that took part in both 2023 and 2024 data collection. Only participants who left their household premises to collect water or do water-related work were included. Once the analytic sample was defined, individual-level data from repeat communities were pooled to compare data from 2023 and 2024 (analysis a1). These comparisons were conducted in the following three steps.

First, we examined whether there were differences in means for data collected in 2023 compared to 2024, regardless of the source type visited. (**Fig 1**, analysis a1.1). Specifically, data from participants from repeat communities were pooled, and mean time, distance, caloric expenditure, and water volume from 2023 were compared with corresponding data from 2024 regardless of water source type. Unequal variance t-tests were conducted for each metric in the Kenya sample. We did not conduct t-tests for the Honduras sample because of the small sample size.

Second, we assessed if there were differences in means for data collected in 2023 compared to 2024, among those who visited unimproved sources only (**Fig 1**, analysis a1.2). This analysis not only examined differences by year among unimproved source users but also provided a point of comparison for the subsequent analysis (a1.3). Specifically, data from participants from repeat communities who visited unimproved sources were pooled, and mean time, distance, caloric expenditure, and water volume from 2023 were compared with like data from those who visited unimproved sources in 2024. Unequal variance t-tests were conducted for each metric in the Kenya sample. We did not conduct t-tests for the Honduras sample because of the small sample size.

Third, we explored if there were differences in means for those who went to an unimproved source in 2023 and those who went to an improved source in 2024 (**Fig 1**, analysis a1.3). Because no participants in community K3 visited improved sources in 2024, women from K1 and K3 were not pooled for this analysis; instead, the sample was restricted to participants from community K1. Mean time, distance, caloric expenditure, and water volume were compared using Mann-Whitney U tests, selected due to the small sample size, which limited the reliability of parametric assumptions. We did not conduct statistical comparisons for the Honduras sample because of the small sample size.

For descriptive purposes, we generated community maps to visually compare participants’ routes for water collection, water source types, and activities conducted in 2023 and 2024.

##### Repeat participant analysis

The analytic sample for this analysis included women who participated in both 2023 and 2024 as repeat participants (**Fig 1**, analysis a2). Participants who left their household premises to collect water were included. We generated maps to visually compare each participant’s routes for water collection, water source types, and activities conducted across the two years. We also graphed the time participants’ journeys took, including start and end times, to identify similarities or differences between 2023 and 2024. We did not conduct statistical comparisons because of the small sample sizes.

#### 2.6.2 Research Question b: Are women’s water collection experiences different among those who collect water from improved sources compared to those who collect water from unimproved sources?

The analytic sample for research question b included communities where some participants went to improved sources and others went to unimproved sources in either 2023 or 2024. Communities in which all participants went to improved sources or all participants went to unimproved sources were excluded, as these communities did not provide within-community variation in source type visited. Participants who left their household premises to collect water or do water-related work were included; those who did not leave their household premises were not included. Once the analytic sample was defined, individual-level data were pooled to compare data from those who went to improved sources to those who went to unimproved sources. These comparisons were conducted in the following three steps.

First, we examined if there were differences in means for those who went to improved versus unimproved sources, regardless of data collection year (**Fig 1**, analysis b1). We pooled individual-level data from 2023 and 2024 and sorted by source type visited. We then compared mean time, distance, caloric expenditure, and water volume between those who visited improved sources to those who visited unimproved sources. Unequal variance t-tests were conducted for each metric in both the Kenya and Honduras samples.

Second, we assessed if there were differences in means for those who went to improved versus unimproved sources in 2023 only (**Fig 1**, analysis b2). The purpose of this analysis was to compare data within the same data collection period to account for seasonal differences between years. We restricted the sample to include participants from 2023 only and again sorted women by source type visited. We then compared mean time, distance, caloric expenditure, and water volume between those who visited improved sources to those who visited unimproved sources. Unequal variance t-tests were conducted for each metric in the Kenya sample. We did not conduct t-tests for the Honduras sample because of the small sample size.

Third, we examined if there were differences in means for those who went to improved versus unimproved sources in 2024 only (**Fig 1**, analysis b3). As above, the purpose of this analysis was to compare data within the same data collection period to account for seasonal differences between years. We restricted the sample to include participants from 2024 only and again sorted women by source type visited. We then compared mean time, distance, caloric expenditure, and water volume between those who visited improved sources to those who visited unimproved sources. Unequal variance t-tests were conducted for each metric in the Kenya sample. We did not conduct t-tests for the Honduras sample because of the small sample size.

For descriptive purposes, community-level maps were first created to visually display similarities or differences in the participants’ routes, water source types, distances, and activities conducted at improved and unimproved water sources. We also produced a graph of mean journey time by water source type to highlight any community-level patterns.

#### 2.6.3 Research Question c: How does women’s water-related work differ between those who use water from on-premises sources compared to those who collect water from off-premises sources?

The analytic sample for research question c included all participants in Honduras in 2024 (**Fig 1**, analysis c). To address this question, we sorted participants into two groups (those who did water work on household premises versus those who fetched water from sources off premises), generated descriptive statistics by group, and determined mean time, caloric expenditure and water volume attained for each group. We did not carry out unequal variance t-tests for time and caloric expenditure because we felt these metrics were not directly comparable given how different the on-premises and off-premises activities were. Unequal variance t-tests were conducted for water volume only as we felt the amount of water handled to be comparable. Quantitative analyses were conducted in Stata (version 19), maps were created with ArcGIS (3.5.0), and graphs were generated in R (2026.01.0+392).

### 2.7 Ethics

The study protocol was reviewed by the Institutional Review Board committee of Emory University, Atlanta, Georgia, USA, which determined that the research was exempt from further review and approval (IRB00005955). The protocol was approved by Universidad Nacional Autónoma de Honduras (CEIFCS-2023-P18) in Honduras; St. Paul’s University - Institutional Scientific Ethics Committee (ERB No. 38), the National Commission for Science, Technology and Innovation (NACOSTI/P/23/27117 and NACOSTI/P/24/37524 in 2024) in Kenya. All participants provided informed consent. Participants in Honduras and Kenya consented verbally, and enumerators signed the consent form to affirm. Additional information regarding the ethical, cultural, and scientific considerations specific to inclusivity in global research is included in the Supporting Information (**S1 Checklist**).

## 3 RESULTS

### 3.1 Sample and participant characteristics

A total of 133 unique women participated in data collection from Kenya and Honduras in 2023 and 2024: 66 in Kenya (18 in 2023, 44 in 2024, 4 in both years) and 67 in Honduras (16 in 2023, 50 in 2024, and 1 in both years). Water collection experiences of women who participated in data collection in Kenya and Honduras in 2023 (e.g., time, distance, energy, weight and volume of water) were described previously [4]. Mirroring our analysis from this previous paper, we describe the water collection experiences of all 2024 participants (n = 133) in **S1 Table**.

To answer research questions a, b, and c, data from 92 of the 133 women were included in analysis (**Table 2**). Specifically, 38 unique women from Kenya were included (14 from 2023, 20 from 2024, and 4 from both years) and 54 unique women from Honduras were included (4 from 2023, 49 from 2024, and 1 who participated both years). Reasons for women being excluded from analysis include: being in a community that was not engaged in both 2023 and 2024 (research question a), being in a community where participants all went to unimproved sources or all went to improved sources, limiting intra-community comparison (research question c), or engaging in water work on-premises (research questions a and b for Honduras only).

**Table 2.** Analytic samples for each research question, with the number of communities and participants included and excluded by country setting and year.

| **Analytic sample for Question a: Are women's water collection experiences different in 2023 compared to 2024?** |  |  |  |  |  |  |  |  |
| --- | --- | --- | --- | --- | --- | --- | --- | --- |
|  | **Kenya** |  |  |  | **Honduras** |  |  |  |
|  | 2023 |  | 2024 |  | 2023 |  | 2024 |  |
|  | Included | Excluded | Included | Excluded | Included | Excluded | Included | Excluded |
| **Communities engaged** | **2** | **4** | **2** | **4** | **1** | **5** | **1** | **5** |
| Improved water source delivered<sup>1</sup> | 0 | 3 | 2 | 1 | 0 | 0 | 0 | 2 |
| **Total observations** | **6** | **16** | **16** | **32** | **2** | **14** | **3** | **48** |
| Visited improved source | 0 | 6 | 5 | 10 | 0 | 2 | 1 | 43 |
| Visited unimproved source | 6 | 10 | 11 | 22 | 2 | 12 | 2 | 5 |
| **Repeat participants** | **4** | — | **4** | — | **1** | — | **1** | — |
| Visited improved source | 0 | — | 2 | — | 0 | — | 0 | — |
| Visited unimproved source | 4 | — | 2 | — | 1 | — | 1 | — |
| **Analytic sample for Question b. Are women's water collection experiences different among those who collect water from improved sources compared to those who collect water from unimproved sources?** |  |  |  |  |  |  |  |  |
|  | **Kenya** |  |  |  | **Honduras** |  |  |  |
|  | 2023 |  | 2024 |  | 2023 |  | 2024 |  |
|  | Included | Excluded | Included | Excluded | Included | Excluded | Included | Excluded |
| **Communities engaged** | **3** | **3** | **2** | **4** | **1** | **5** | **3** | **3** |
| Improved water source delivered<sup>1</sup> | 3 | 0 | 1 | 2 | 0 | 0 | 0 | 2 |
| **Total observations** | **12** | **10** | **16** | **32** | **3** | **13** | **10** | **41** |
| Visited improved source | 6 | 0 | 7 | 8 | 1 | 1 | 4 | 40 |
| Visited unimproved source | 6 | 10 | 9 | 24 | 2 | 12 | 6 | 1 |
| **Analytic sample for Question c. How does women's water-related work differ between those who use water from on premises sources compared to those who use water from off-premises sources?** |  |  |  |  |  |  |  |  |
|  | **Kenya** |  |  |  | **Honduras** |  |  |  |
|  | 2023 |  | 2024 |  | 2023 |  | 2024 |  |
|  | Included | Excluded | Included | Excluded | Included | Excluded | Included | Excluded |
| **Communities Engaged** | — | — | — | — | — | — | **6** | **0** |
| Improved water access delivered | — | — | — | — | — | — | 2 | 0 |
| **Total observations** | — | — | — | — | — | — | **50** | **0** |
| On-premises water work | — | — | — | — | — | — | 40 | 0 |
| Off-premises journey | — | — | — | — | — | — | 10 | 0 |

<sup>1</sup>Improved water source delivered by World Vision prior to data collection. Some participants had access to other improved water sources.

The following descriptive statistics are for participants included in analysis. In Kenya, the mean age of participants was 34 years, and the mean household size was 6.2 people (**Table 3**). The majority of women reported never attending school (88%). Most women also reported not engaging in economic activities in the last 30 days in both 2023 (78%) and 2024 (75%). From the survey, most participants reported using unimproved sources for their primary source of drinking water, and the proportion that reported using an unimproved source was lower in 2023 (71%: unimproved dug well, unprotected spring) than in 2024 (92%: unprotected dug well, surface water). Participants in 2024 estimated that the mean total number of trips to the water source visited on the day of data collection was 1.6 times, with an estimated water labor time of 102.7 min for the day. Additionally, the participant-estimated time to manage water at home was 46.4 min. For the demographic and water source characteristics of all Kenya women who participated in data collection in 2024 (n = 48), see **S2 Table.**

**Table 3.**
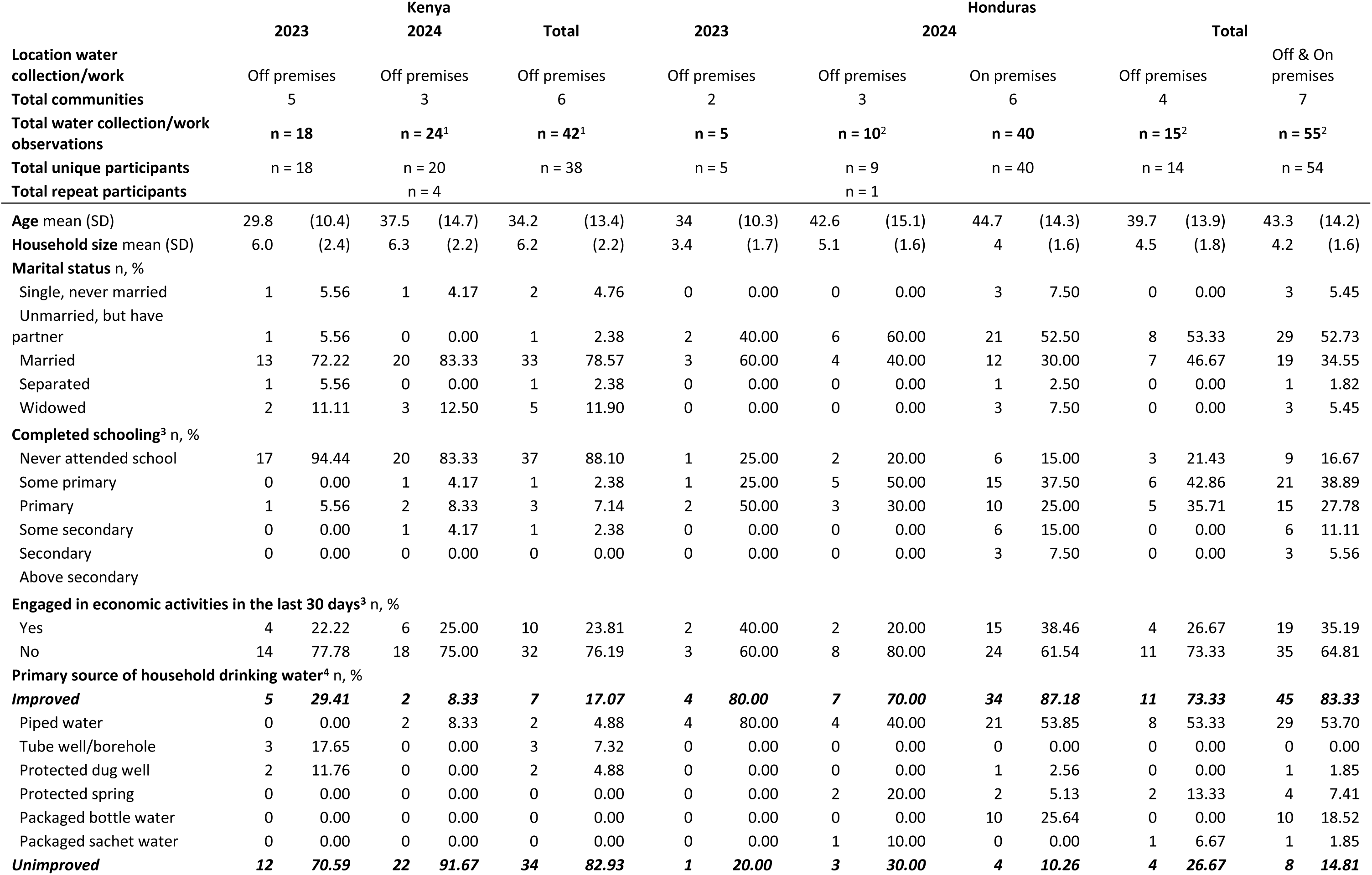
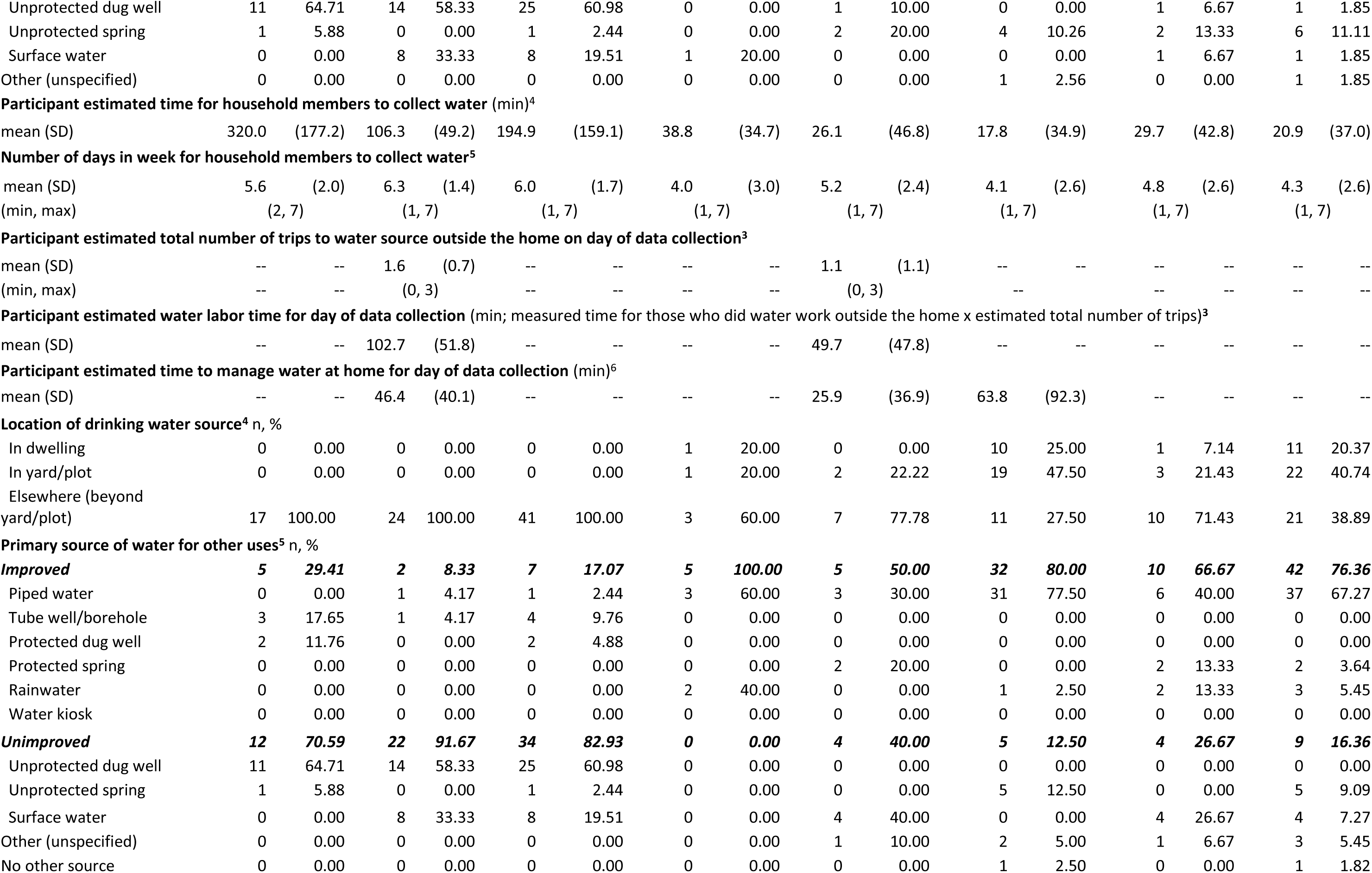
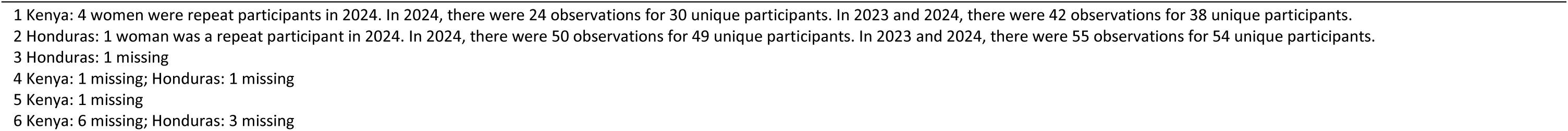
Demographic and water characteristics of participants included in analysis, by country and year of data collection.

In Honduras, the mean age of participants was 43 years old, and the mean household size was 4.2 people (**Table 3**). Most women had a primary education or less (83%). The majority of women reported that they had not participated in economic activities in the last 30 days in both 2023 (60%) and 2024 (65%). In the survey, most participants said that they used improved sources for their primary source of drinking water, and the proportions were similar in 2023 (80%: piped water) and 2024 (85%: piped water, packaged bottled water, protected spring, packaged sachet water, protected dug well). In 2024, participants who went off-premises during data collection estimated making an average of 1.1 total trips to the water source on the day of data collection, with an estimated water labor time for the day of 49.7 min. Additionally, the participant-estimated time required to manage water at home was 25.9 min for women who went off-premises and 63.8 min for women who stayed on-premises.

### 3.2 Findings for research question a: Are women’s water collection experiences different in 2023 compared to 2024?

Two communities in Kenya, K1 and K3, and one community in Honduras, H1, were engaged in data collection in both 2023 and in 2024 and included in analysis (‘repeat communities’). Four women in Kenya and one woman in Honduras participated both years (‘repeat participants’).

#### 3.2.1 Kenya repeat community results

In community K1 in 2023, all participants (n = 3) went to unimproved sources (unprotected dug wells), whereas in 2024, five participants went to an improved source (public piped water; constructed after 2023 data collection), and three participants went to unprotected dug wells (**Fig 2**). In 2023, activities included digging unprotected dug wells and collecting water. In 2024, activities included these same tasks, along with bathing self, bathing children, and collecting firewood; however, women who used the improved piped water did not need to dig, unlike those using unprotected dug wells.

**Fig 2.**
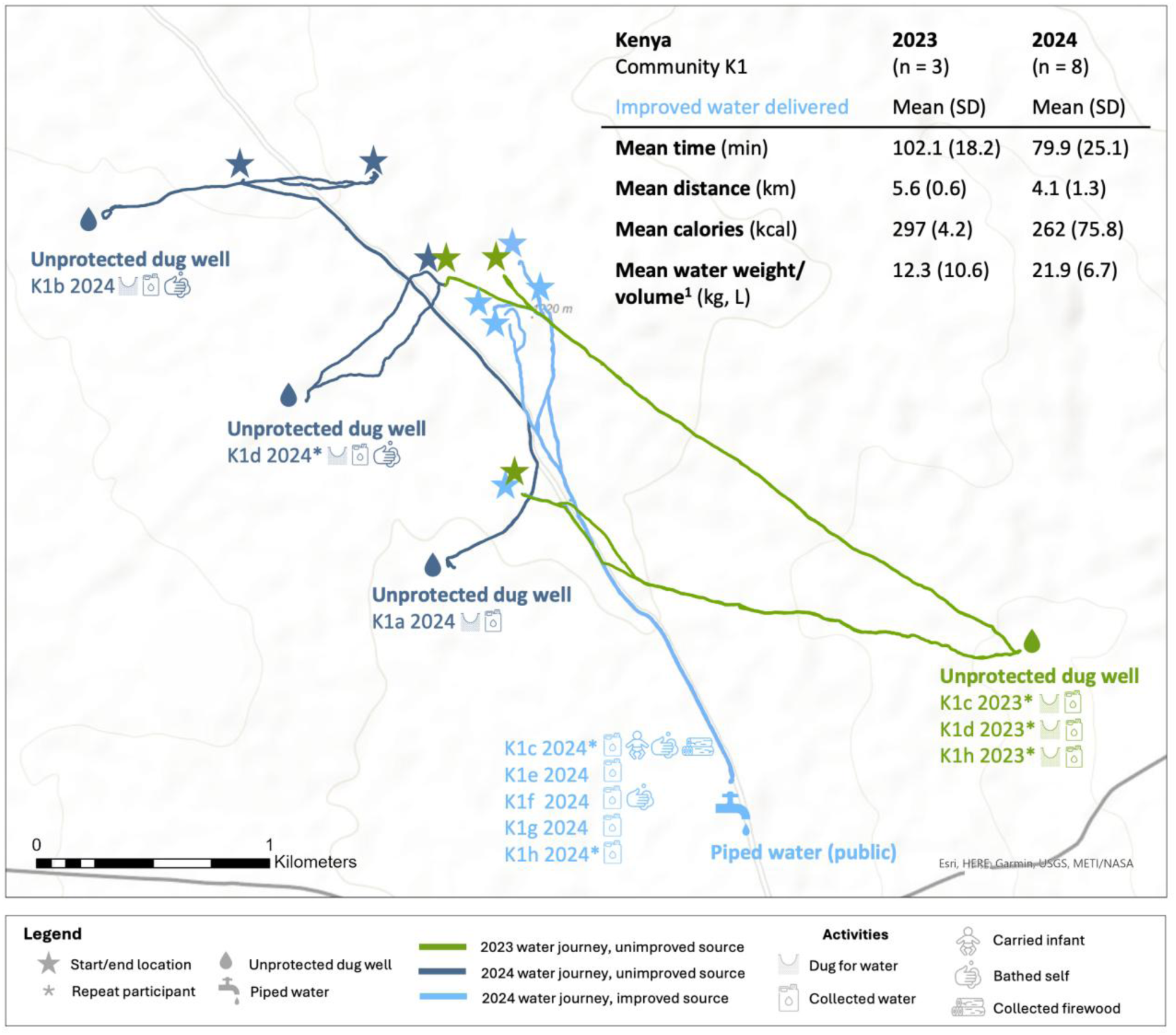
**Map of round-trip routes to water sources and activities conducted in 2023 and 2024, Kenya community K1**. ^1^ 2024: 1 missing water weight/volume

In community K3 in 2023, all participants (n = 3) went to unprotected dug wells, and in 2024, all participants (n = 8) went to unimproved surface water sources (dams) (**Fig 3**). For both years, all participants collected water, although those who used surface water in 2024 did not need to dig to reach water, in contrast to the women who used unprotected dug wells for water.

**Fig 3.**
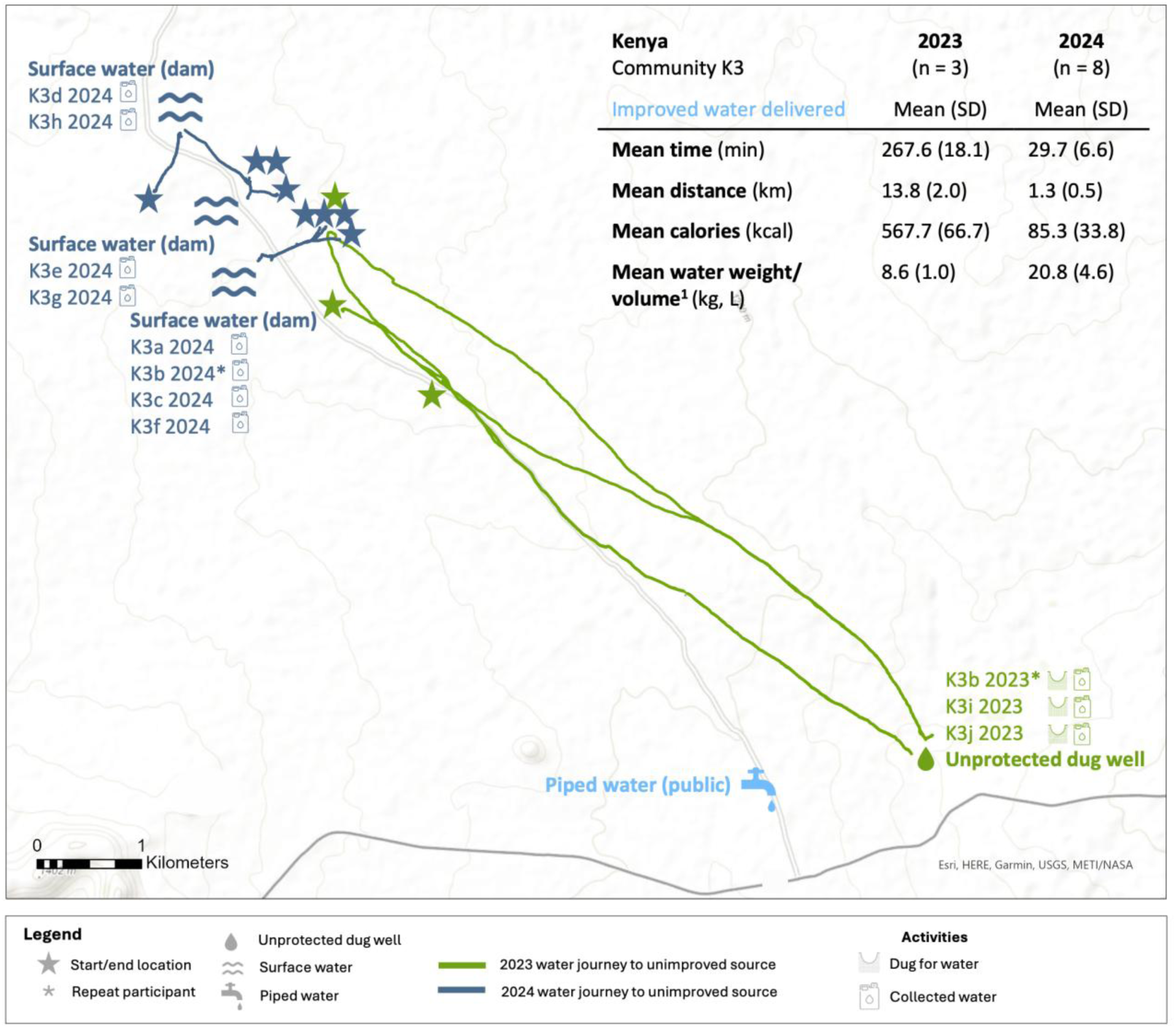
**Map of round-trip routes to water sources and activities conducted in 2023 and 2024, Kenya community K3.** ^1^ 2024: 1 missing water weight/volume

In pooled analyses comparing data from those in 2023 to those in 2024 regardless of source type visited (analysis a1.1), we found that journeys to sources in 2024 took significantly less time, distance, and caloric expenditure than in 2023 (time: 54.8 vs 184.8 min, t = 3.4, p=.017; distance: 2.7 vs 9.7 km, t = 3.6, p=.014; caloric expenditure: 173.6 vs 432.5 kcal, t = 3.8, p=.007) (**Table 4**). Women also collected significantly greater volumes of water during the single trip in 2024 than in 2023 (water volume: 21.4 vs 10.8 L, t = −2.8, p=.035; water volume/household member: 3.9 vs 1.7 L/person, t = −2.9, p =.011).

**Table 4.**
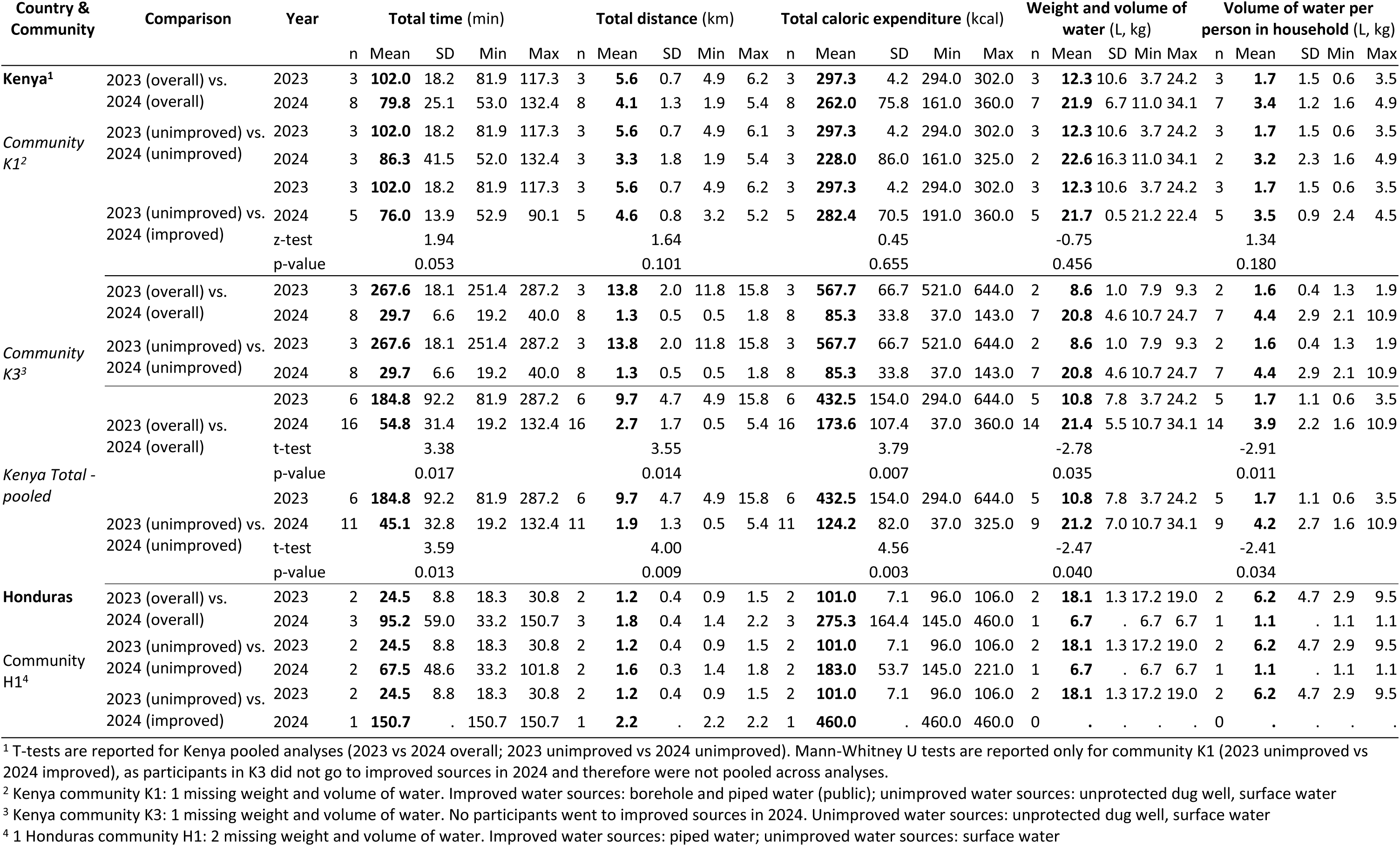
Comparison of repeat communities in Kenya and Honduras in 2023 and 2024.

Similarly, in pooled analyses comparing data from those who went to unimproved sources in 2023 to 2024 (analysis a1.2), we found that journeys to sources in 2024 required significantly less time, distance, and caloric expenditure than in 2023 (time: 45.1 vs 184.8 min, t = 3.6, p=.013; distance: 1.9 vs 9.7 km, t = 4.0, p=.009; caloric expenditure: 124.2 vs. 432.5 kcal, t = 4.6, p=.003) (**Table 4**). Again, participants collected significantly greater volumes of water during the single trip in 2024 than in 2023 (water volume: 21.2 vs 10.8 L, t = −2.5, p=.040; water volume/household member: 1.7 vs 4.2 L/person, t = −2.4, p=.034).

In pooled analyses comparing data from those who went to unimproved sources in 2023 to those who went to improved sources in 2024 (analysis a1.3), there were no significant differences in time, distance, caloric expenditure, or water volume (time: 102.0 vs 76.0 min, z = 1.9, p=.053; distance: 5.6 vs 4.6 km, z=1.6, p=.101; calories: 297.3 vs 282.4 kcal, z = 0.5, p=.655; water volume: 12.3 vs 21.7 L, z = −0.8, p=.456; water volume/household member: 1.7 vs 3.5 L/person; z = 1.3, p=.180) (**Table 4**).

#### 3.2.2 Honduras repeat community results

In Honduras, one community (H1) was engaged in data collection in both 2023 and 2024, and data was collected from five participants (2 in 2023, 3 in 2024) (**Fig 4**). In 2023, both participants went to surface water (river), and in 2024, two participants went to surface water (river), and one went to piped water at neighbor’s household. Activities in 2023 were limited to collecting water, whereas activities in 2024 included collecting water, bathing self, bathing child, and washing clothes.

**Fig 4.**
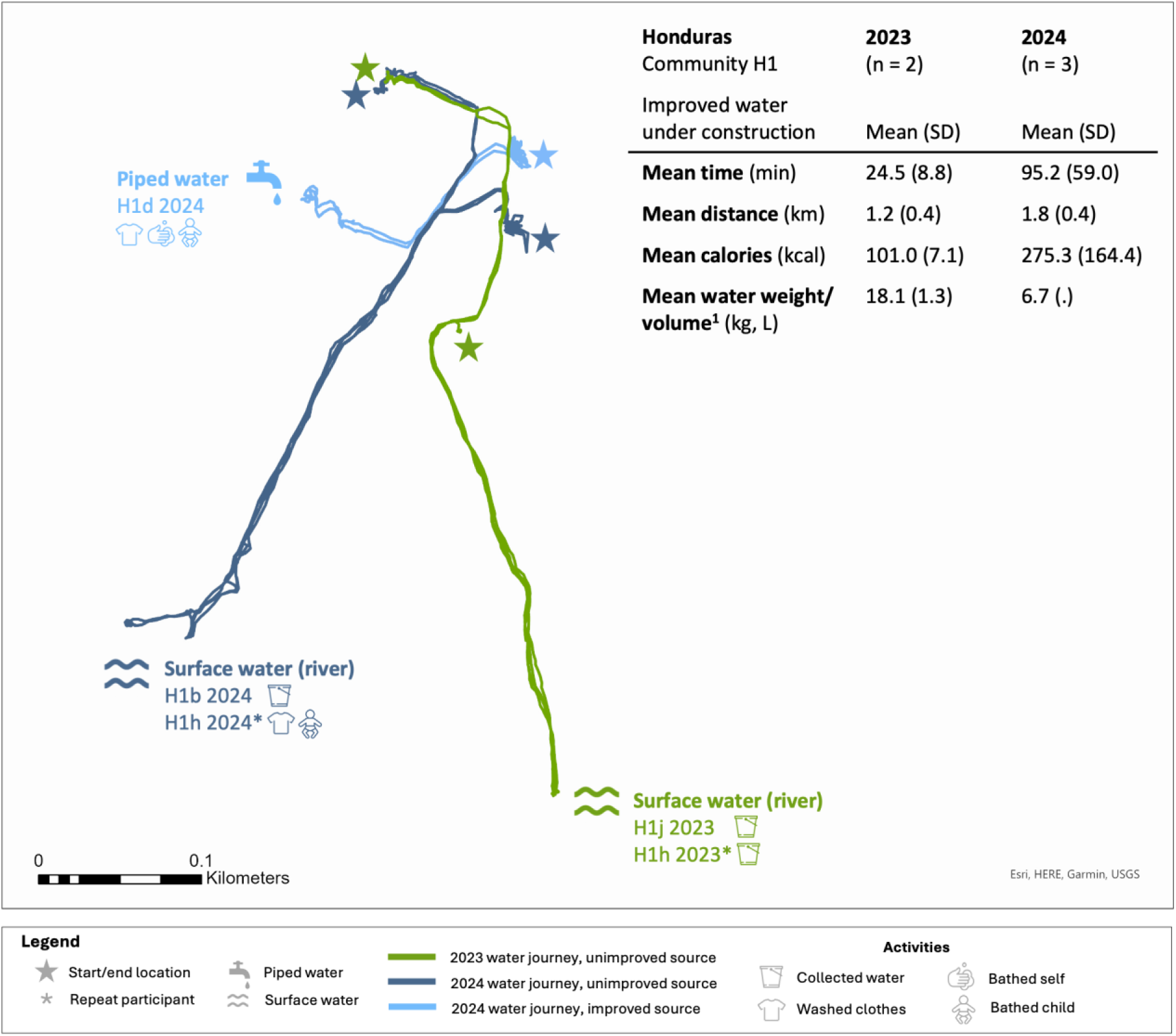
**Map of round-trip routes to water sources and activities conducted in 2023 and 2024, Honduras community H1.** ^1^ 2024: 2 missing water weight/volume

We did not conduct pooled analyses comparing data from those in H1 because of the small sample size.

#### 3.2.3 Kenya and Honduras repeat participant results

Only 5 participants (4 in Kenya, 1 in Honduras) participated in data collection in both 2023 and 2024, limiting analyses. Among the four repeat participants in Kenya, two walked to unimproved sources in 2023 and used improved sources in 2024, and two used unimproved sources both years. The Honduras repeat participant visited an unimproved source in both 2023 and 2024.

Additional information can be found in the supplement, including descriptive data (**S3 Table**), maps of repeat participant journeys (**S1 & S2 Figs**) and a bar graph showing total time, timing during the day, and water source type visited of repeat participants (**S3 Fig**).

### 3.3 Findings for research question b: Are women’s water collection experiences different among those who collect water from improved sources compared to those who collect water from unimproved sources?

Five communities in Kenya and four communities in Honduras had participants who went to a mix of improved and unimproved sources and were eligible for analysis.

#### 3.3.1 Kenya results

In Kenya, 13 participants (6 in 2023; 7 in 2024) went to improved sources (borehole, piped water) and 15 (6 in 2023; 9 in 2024) went to unimproved sources (unprotected dug well, surface water) (**Figs 5 & 6, S4 Fig**). Women in 2024 used more varied source types and locations than participants used in 2023. For example, participants went to multiple locations to access water from unprotected dug wells. Across both years, all participants collected water. Other activities at improved sources or along the trip included bathing self and children and collecting firewood. At unimproved sources, women performed these same activities, with the addition of digging to access water from the unprotected dug wells and washing clothes.

**Fig 5.**
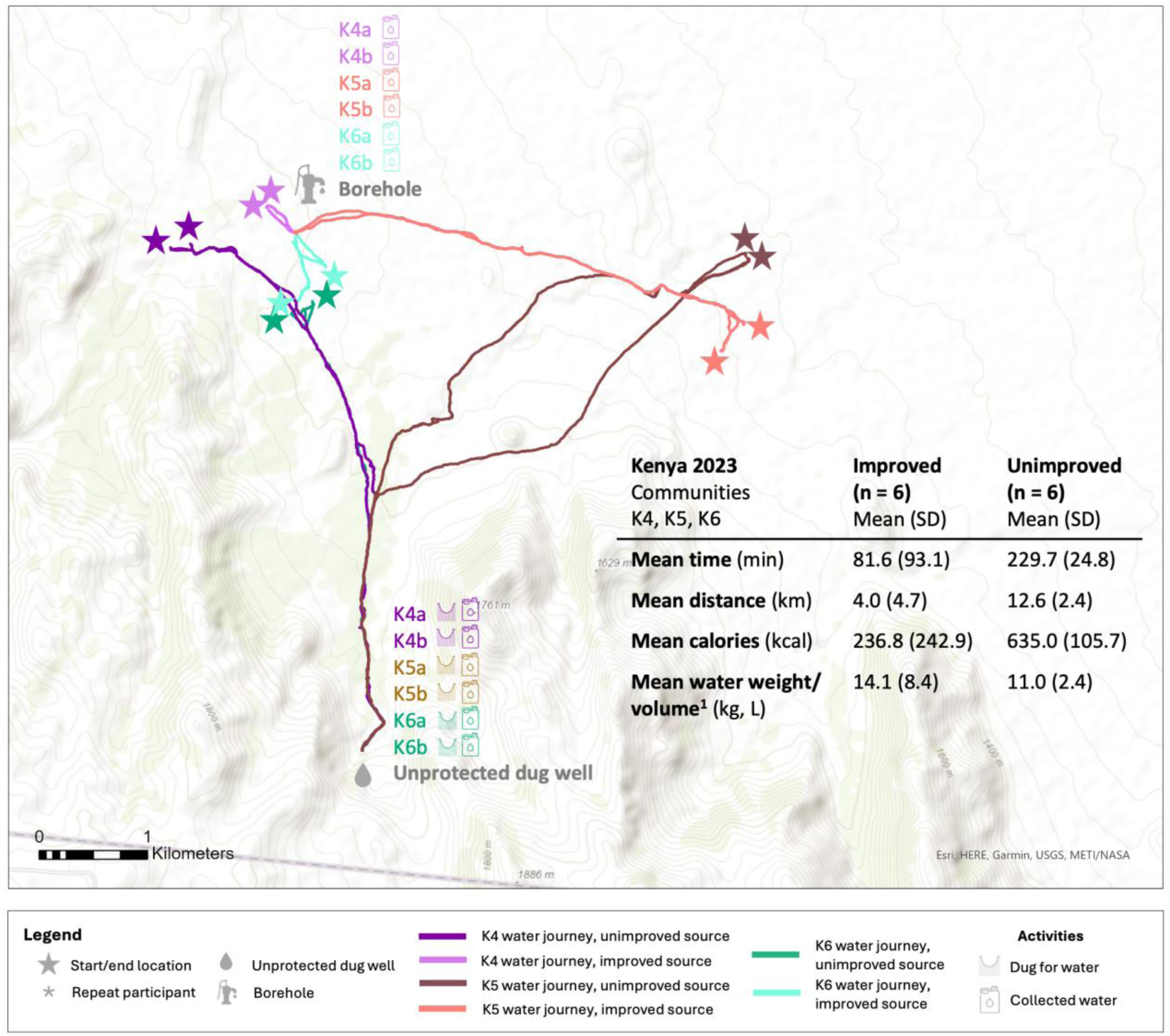
**Map of round-trip routes to water sources and activities conducted in 2023, Kenya communities K4, K5, K6.** ^1^ Unimproved: 2 missing water weight/volume

**Fig 6.**
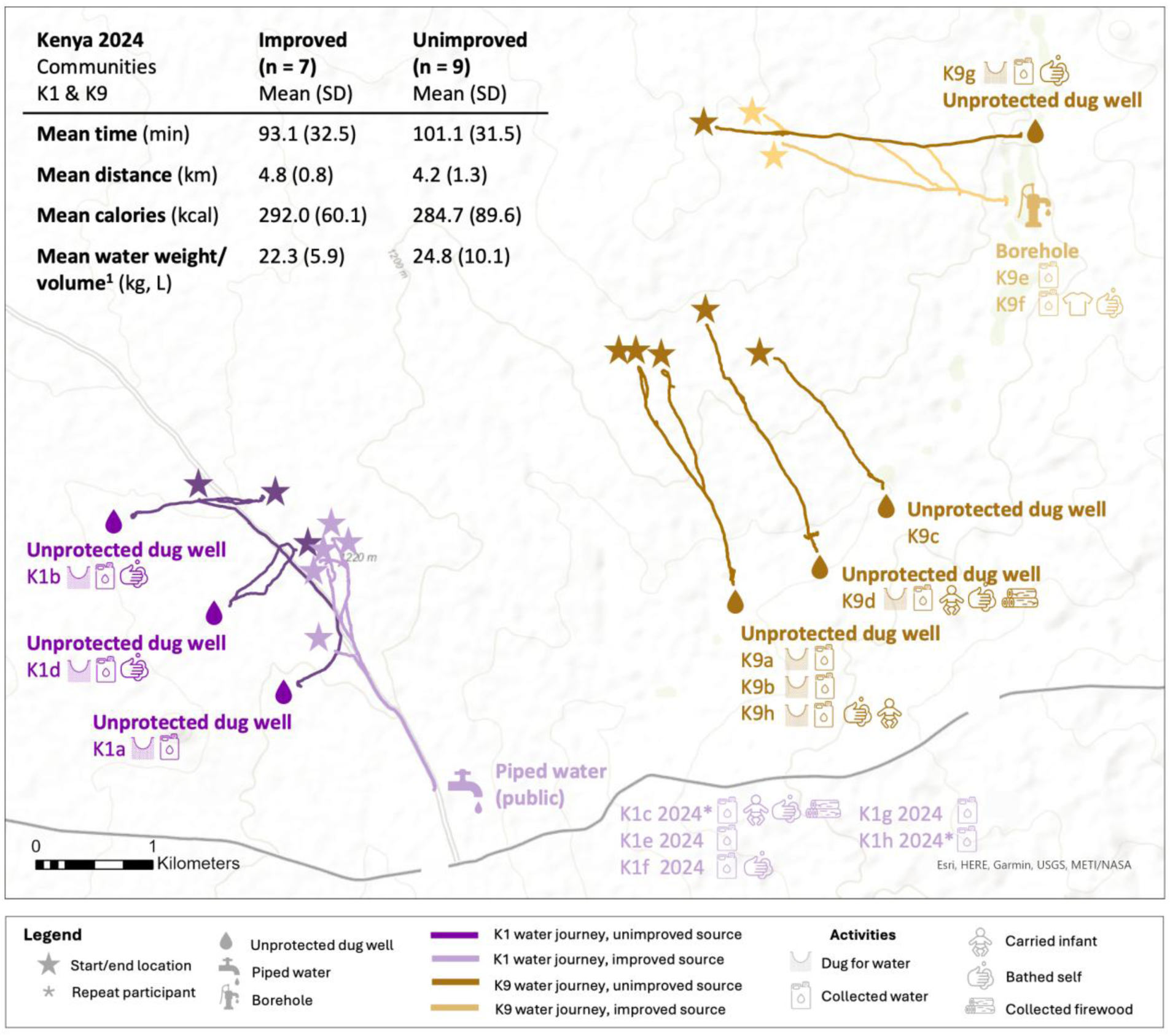
**Map of round-trip routes to water sources and activities conducted in 2024, Kenya communities K1 and K9**. ^1^ Unimproved: 1 missing water weight/volume

In pooled analyses of data from both years (analysis b1), journeys to improved sources took significantly less time, distance, and energy expenditure than journeys to unimproved sources (time: 87.8 vs 152.6 min, t = −2.5, p=.018; distance 4.4 vs 7.5 km, t = −2.1, p=.043; caloric expenditure: 266.5 vs 424.8 kcal, t = −2.3, p=.030) (**Table 5**). Water volume and water volume per household member did not differ by source type visited (volume: 18.5 vs 20.2 L, t = −0.4, p=.666; volume per household member: 3.2 vs 3.5 L/person, t = −0.4, p=.666).

**Table 5.**
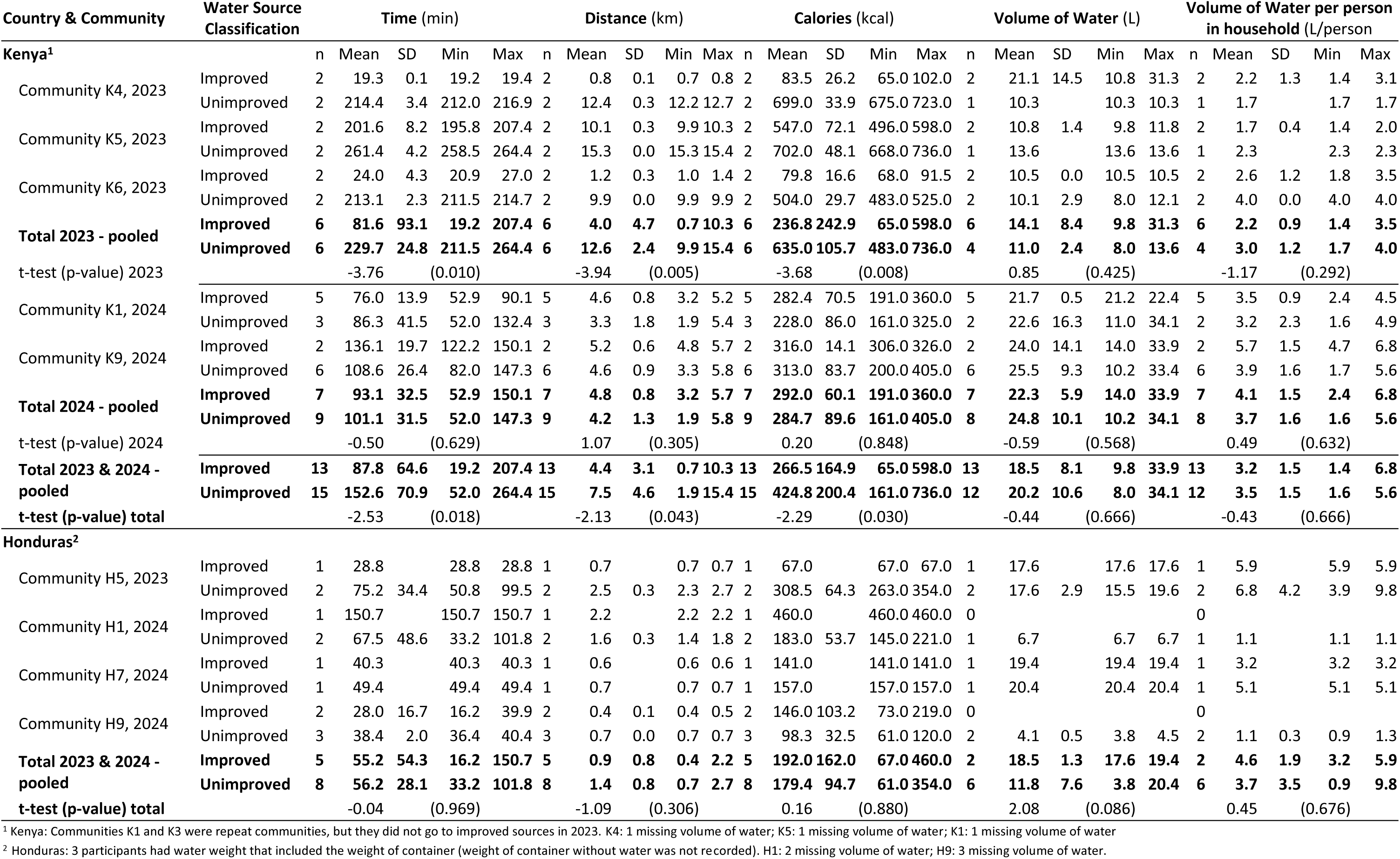
Comparison of communities where participants went to improved and unimproved sources at the same time point, by country.

Similarly, in pooled analyses of data from 2023 alone (analysis b2), journeys to improved sources required significantly less time, distance, and energy expenditure than journeys to unimproved sources (time: 81.6 vs 229.7 min, t = −3.8, p=.010; distance: 4.0 vs 12.6 km, t = −3.9, p=.005; caloric expenditure: 236.8 vs 635.0 kcal, t = −3.7, p=.008) (**Table 5**). Again, there were no differences in water volume by source type visited (volume: 14.1 vs 11.0 L, t = 0.9, p=.425; volume per household member: 2.2 vs 3.0 L/person, t = −1.2, p=.292).

However, in pooled analyses from 2024 alone (analysis b3), no significant differences were observed between journeys to improved sources compared to unimproved sources for time, distance, energy expenditure, or water volume (time: 93.1 vs 101.1 min, t = −0.5, p=.629; distance: 4.8 vs 4.2 km, t = 1.1, p=.305; caloric expenditure: 292.0 vs 284.7 kcal, t = 0.2, p=.848; volume: 22.3 vs 24.8 L, t = −0.59, p=.568; volume per household member: 4.1 vs 3.7 L/person, t = 0.49, p=.632) (**Table 5**).

#### 3.3.2 Honduras results

In Honduras, five participants (1 in 2023; 4 in 2024) visited improved sources (piped water, protected spring) and eight (2 in 2023; 6 in 2024) went to unimproved sources (surface water, unprotected spring) (**Fig 7**). Activities at improved and unimproved source types were largely similar and included collecting water, washing clothes, and bathing self and children. Not all women collected water during their journey. Other activities included washing dishes and cleaning the area around the water source.

**Fig 7.**
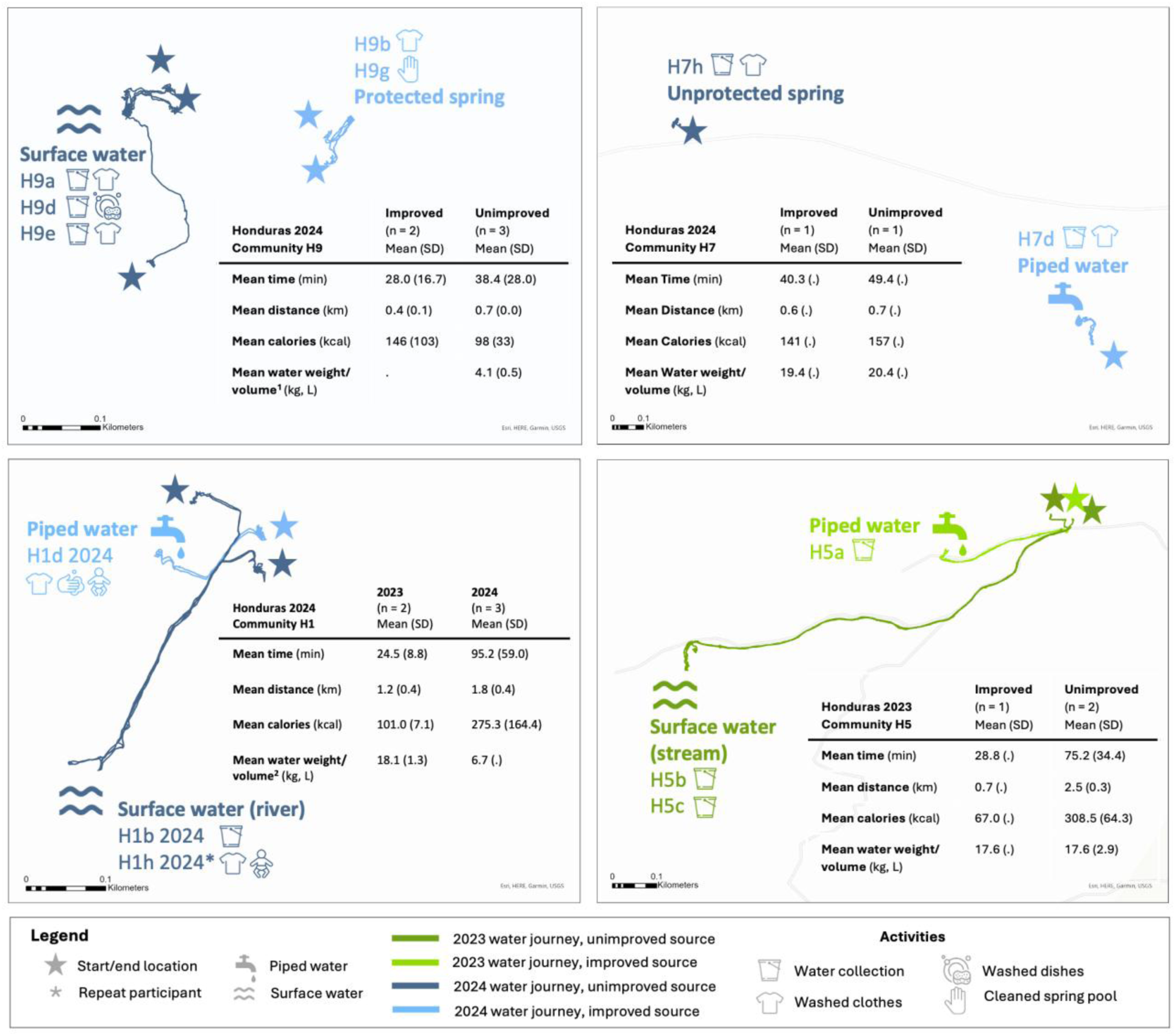
**Maps of round-trip routes to water sources and activities conducted in 2024, Honduras communities H9, H7, H1 and H5**. ^1^ H9 improved: 2 missing water weight/volume; unimproved: 1 missing water weight/volume ^2^ H1 improved: 1 missing water weight/volume; unimproved: 1 missing water weight/volume

In pooled analyses of data from both years (analysis b1), no significant differences were observed between journeys to improved compared to unimproved sources for time, distance, caloric expenditure, and water volume (time: 55.2 vs 56.2 min, t = −0.04, p=.969; distance: 0.9 vs 1.4 km, t = −1.1, p=.306; caloric expenditure: 192.0 vs 179.4 kcal, t = 0.2, p=.880; volume: 18.5 vs 11.8 L, t = 2.1, p=.086; volume per household member: 4.6 vs 3.7 L/person, t = 0.5, p=.676) (**Table 5**).

### 3.4 Research question c: How does women’s water-related work differ between those who use water from on-premises sources compared to those who collect water from off-premises sources?

Among participants in 2024, 20% (n = 10) collected water from off-premises water sources and 80% (n = 40) remained on-premises to carry out water-related work, either in their home or yard (**Table 6**). Participants in Honduras who remained on-premises during 2024 data collection engaged in a range of water-related activities. Women collected water from *pila* water storage containers, washed and hung clothes, prepared food (e.g., washed cooked corn, made a bottle for an infant), cleaned (e.g., household surfaces, toilets, *pilas*), bathed themselves and their children, watered household plants, and purchased water from delivery trucks. Participants who traveled off premises for water engaged in a similar but narrower set of activities, including collecting water, washing clothes, bathing themselves and a child, washing dishes, and cleaning the area around a water source.

**Table 6.**
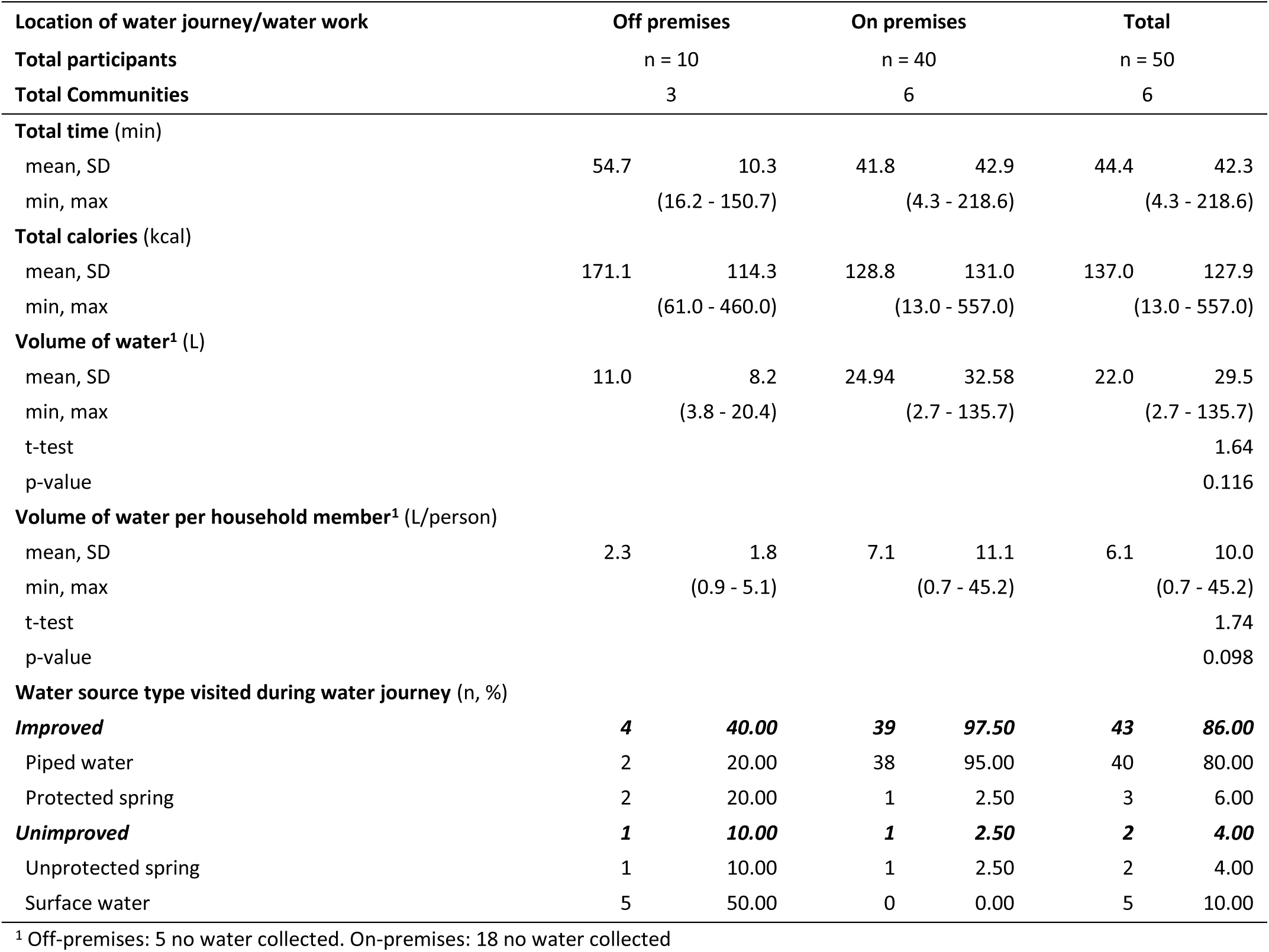
Water-related work experiences among Honduras participants on and off-premises in 2024.

No significant differences were observed between off-premises journeys compared to on-premises water collection for mean water volume collected (volume: 11.0 vs 25.0 L, t = 1.6, p=.116; volume per household member: 2.3 vs 7.1 L/person, t = 1.7, p=.098). For on-premises observations, water was typically collected from *pila* water storage containers. Mean total time and caloric expenditure appeared to be greater among participants who went outside of their household compound (54.7 min, 171.1 kcal) compared to women who remained on premises (41.8 min, 128.0 kcal). As noted above, no tests of comparison were conducted for these two metrics.

## 4 DISCUSSION

Our study compared women’s water collection and water work experiences in rural Kenya and Honduras between different time points (2023 and 2024), improved and unimproved water source types, and on-and off-premises sources. Women’s experiences varied by year and setting, with changes shaped by source type, seasonal conditions, and source location. In Kenya, mean water collection time, distance, and caloric expenditure were significantly lower and water volume was significantly higher in 2024, when heavy rains enabled water collection from proximate sources, compared to 2023, when drought limited access. When comparing source types during the 2023 drought, journeys to improved sources took significantly less time and energy and covered shorter distances than journeys to unimproved sources; however, no significant differences were observed during the rainy conditions of 2024, when unimproved sources became closer and more accessible than improved sources. In Honduras, off-premises water collection burdens did not differ significantly by year or source type. Notably, women with on-premises water access still expended considerable time and energy performing water work within their household compounds. These findings further highlight the demanding nature of water collection and related work and the influence of environmental conditions, even when improved sources are available.

In both Kenya and Honduras, water collection and related work continued to place substantial time demands on women, underscoring that burdens persist, especially when water access remains off premises. Previous studies have reported reductions in time following water supply interventions; however, for most studies, collection times to new sources were shorter and largely under 30 minutes [21–23, 46–50]. An exception is a study in southern Kenya, where women reported that their roundtrip time to new public sources was reduced but still took over 1.5 hours during the dry season [48]. As we have learned from our study, new improved sources may be closer than unimproved sources during dry seasons but still take substantial time and may be less convenient than unimproved sources in rainy seasons. Further, in a companion study from our team that assessed how women in Kenya and Honduras spent time over a 24-hour period, participants in communities that had received improved water infrastructure were not found to spend any less time collecting water compared to those who had not yet received improvements [51]. These findings demonstrate that provision of water does not guarantee the elimination of burdens and underscore the need to prioritize proximity and accessibility alongside water quality, reliability, and sufficient quantity in infrastructure provision. Ensuring household-level water access, where feasible, could more directly reduce women’s physical and time burdens [52]. Evaluations of water infrastructure should include assessments of water collection burden, and recently identified priority gender-specific indicators for WASH under Sustainable Development Goals (SDG) 6.1 and 6.2 offer measures for doing so [53].

Our findings in rural Kenya add to the evidence that seasonal and climatic variation can play a critical role in women’s water source selection [28, 54–56], highlighting the need for implementers to be attentive to seasonal (and potential climatic) changes if infrastructure is intended to be used year-round. Improved water sources in Kenya appeared to reduce water collection burdens, but these changes were most pronounced when comparing women from the same communities who went to different source types (improved and unimproved) during a period of drought in 2023. During this drought, water was difficult to retrieve from unimproved sources, requiring women to travel extremely long distances and dig holes in riverbeds to access it. Those with access to improved sources traveled less far and did not have to do arduous digging labor at the sources. In contrast, when heavy rain increased surface and groundwater availability in 2024, differences in time, distance, and energy between improved and unimproved sources narrowed. As our maps show, in one community (K3), surface water sources were close to women’s homes — much closer than the improved public piped supply (**Fig 7**). As a result, all women in this community collected from these proximate, and therefore more convenient, surface water locations. In the other repeat community (K1), while over half of the participants went to the improved source, not all did; some chose to collect water from nearby dug wells that were closer to their homes. Our findings are consistent with an earlier study in southern Kenya, where trips were shorter in the wet season than in the dry season [48] and with findings from a study in rural Zambia, where women reported using shallow dug wells during the rainy season because boreholes were farther away [55]. In addition to water availability, seasonality can also influence people’s willingness to pay for water, their perception of water, and the appearance of water, all of which may affect women’s water source selection even when an improved source is available [54, 57–60]. Because of this, water programs should integrate water treatment in addition to improving access, particularly when surface water is the more convenient option, though treatment methods promoted should not further burden women [61–63].

More broadly, our results illustrate how environmental variability, whether expected seasonal transition or unexpected climate-driven events, can influence women’s access to and selection of water sources. We intentionally collected data at the same time of year to capture comparable seasons, but unexpected rainfall created different conditions, highlighting how women adapt to environmental shifts. While limited, our results are stark given the recent estimates that warming from climate change by 2025 may increase women’s time spent on water collection by 30% globally and 100% regionally [64]. The findings from Kenya show that changes in the environment can limit or broaden water source options. Women may may not select an available improved source if more convenient options become available, even if they are less safe. Multiple water source use may help diversify risk and strengthen resilience to climate change, though additional sources may be unimproved [65]. As climate change threatens more frequent and intense stresses, water initiatives must not only integrate climate resilience into planning and monitoring [66], but ensure that women’s needs, responsibilities, and voices are integrated [67].

Our findings also underscore the need for research and monitoring to extend beyond water collection from primary drinking water sources, as this practice may overestimate basic and safely managed access in settings where households rely on multiple water sources [54]. Multiple water source use is common globally, including among households with improved water, and is influenced by what the water is for (e.g., drinking, cooking, bathing), water characteristics (e.g., perceived cleanliness, color, taste), source characteristics (e.g., distance, intermittency, cost, privacy), and cyclical patterns like season and time of day [4, 28, 54, 68]. For example, women in India and in our earlier study in Guatemala used rivers for washing clothes because water flow helped the activity, even though they had access to other sources [4, 28]. In this study, data collection was conducted with women for one day. However, because water needs can vary by day, participants who used improved sources during data collection may rely on unimproved sources at other times for different purposes, and vice versa, meaning that single-day assessments may inadequately account for the diversity of water source use. A study by Casas et al. (2026) approached this limitation by using high-frequency survey data [68]. Future research could similarly strengthen measurement of multiple water source use by applying our approach over periods longer than a day (e.g., one week) to better understand, particularly before and after improved water installation.

Regardless of water source type, participants in Kenya generally collected more water when water sources were closer; however, when accounting for household size, the volume collected remained insufficient for their needs, indicating potential health implications. For example, Kenyan women in repeat communities increased their average volume collected (11 L to 21 L) as their average distance decreased (10 km to 3 km), but their total water gain translated to less than 4 L per household member, levels that remain critically low. Although improved water source types did enable collection of greater water volumes during 2023 drought conditions, this advantage was not consistent across analyses and likely reflects the physical constraints of water carriage, since women typically carried a standard 20 L jerry can regardless of source type. These findings are important considering a WHO report that defined water access below 5.3 L per person per day as a very high health risk [69] and that an average adult woman requires 2.7 L per day for drinking alone [70]. Making multiple trips per day would increase per-household-member water volume, however, doing so would require additional time and energy from women — and water for their own hydration.

Findings from Honduras show that water work done at home is still demanding even when women have household access, highlighting the need for further research on the scope and burden of water work after water is secured. This study focused primarily on assessing water collection and water-related work among those who had to go to water sources beyond the household, but that scope is limited.

Among the women in Honduras who did water-related work using on-premises sources, we found that they spent an average of 42 min, expended 129 kcal, and moved 25 L of water during the single data collection period. While we lack more granular data on the specific water-related activities performed and the time, energy, and water quantities spent on each, our tracking provides validation that water work does not end once water enters the home. Instead, these findings suggest that previous assessments of water burden, including our own, have been incomplete by focusing only on water collection and overlooking the continued demands on women’s time and energy after water is secured. Further, women also expend cognitive and emotional labor managing the water they do have, work which is invisible and hard to measure [12]. Other methodologies, such as semi-structured observations to assess household water work (leveraging, for example, other semi-structured household behavior observations using customized LiveTrak software [71]), could enable greater understanding of activities in such settings, including what water work is performed, the frequency and duration of water work activities, and who performs them.

## 5 STRENGTHS & LIMITATIONS

This study had a number of strengths. Its longitudinal design allowed comparisons across the same communities at the same time of year, and the use of wearable GPS technologies provided detailed measurements of women’s journeys. Partnering with our Kenya data collection team both years further supported comparability. Participants selected where they would go during data collection, which reflected their decision-making and allowed us to record journeys that aligned with their needs and routines. Additionally, leveraging watch data to create maps enabled a visual understanding of water collection routes for participants and within communities, illuminating trends that are not evident from tabulating data alone.

This work also has limitations. Community sampling was purposive and individual sampling was by convenience for those who were not repeat participants, therefore, the results may not be representative and are not generalizable. The study also did not include participants under 18, whose water collection experiences may differ from those of adults. It was difficult to locate repeat participants, resulting in a smaller than expected sample. While participants chose their own water source destinations, the data collection teams’ presence may have influenced their selection, pace, and activities during their journeys, and thus the data collected.

As we captured only one water collection observation per participant during data collection periods, we may have under-estimated total daily burden because some women may have taken multiple trips or used different sources across one day. To estimate water collection time for the day, we multiplied each participant’s estimated number of daily trips by the measured time of their observed journey; however, this approach may have under- or over-estimated total time and labor. Further research that records all journeys for one day, or over the course of a week, could more accurately characterize total time, labor, and energy, and could be particularly informative if done before and after improved water provision to see if women’s burdens are reduced. Similarly, further research could be conducted on the water work completed within the household.

Caloric expenditure estimates were modeled by the watches, and because watches were worn for only one journey per person, the accuracy of these estimates may be less accurate than if the watches were worn over longer periods. Watches recorded each journey as a single, continuous experience, which limited us from separating the time and energy spent on specific activities like walking to the source or washing clothes; future work could assess the specific contributions of activities. The small sample size limited the statistical power of our comparisons and constrained our ability to examine variation by life stage (e.g., age, marital status).

## 6 CONCLUSION

Our findings highlight how women’s daily lives continue to be shaped by the effort required for water collection and related work, particularly in areas where improved water infrastructure exists but is off premises. These results reinforce that water access is not just about the source type, but about proximity to households, seasonal changes, and the range of household needs women must meet.

Improved water sources remain valuable and can provide benefits to women by reducing time and energy even if not in the home, particularly during dry seasons. Still, while reductions in labor are positive, efforts should strive to eliminate water collection labor as a top priority. Future initiatives can strengthen their impact by involving women in source placement, engaging them to understand how and why they choose particular sources, and planning for seasonal variability and climate resilience. Ultimately, eliminating women’s water labor requires household-level access to water that is affordable, high-quality, and available at all times during the day and year and for all purposes. Methods such as those used in this study and in our companion research [4, 12, 51] can be used to assess the extent to which water interventions alleviate these burdens and support women’s health and well-being.

## Supporting information

S1 Tools

## Data Availability

Add data produced are available online at

https://doi.org/10.6084/m9.figshare.c.8506935

## ACKNOWLEDGEMENTS

We are deeply grateful to the participants for their time and for sharing their experiences. We also extend our thanks to our partners and data collection teams whose collaboration made this research possible. From the Universidad Nacional Autónoma de Honduras: Rosaura Suyapa Rodriguez Funez, Ashly Michelle Méndez, Nastin Nahomy Tilguant Ávila, Brenda Paola Cruz, Yesi Karolina Murillo Vega, Jorge Luis Cálix Barahona, Dulce Milagro Sevilla Sosa, Dulce María Sevilla Sosa, Wilber Adonay Ávila Valladares, Néstor Fabricio Dávila González, and Pedro Alejandro Hernández. From the Edwin Roldan Medina Lopez Research Group: Gladys Rosario Alvarez Montoya, Ruth Avigail Montalban, Ana Carolina Padilla, Luis Antonio Zelaya, Marta Ramos, Pedro Martinez, Salomon Perdomo, Sobeyda Rodriguez, Varinia Dominguez, Mayra Portillo, Maira Velasquez, Luisa Galindo, and Hernan Lanza. From World Vision Honduras: Jazmina Nohemí Irías, Ana Lili Burgoos, Carlos Parrales. From St. Paul’s University: Lesoito Jeremiah Christopher, Leshaule Benedicto Alois, Patricia Lekiliyo, Lechalote Ben Ljania, Nasieku Lolchura, Lekalantula Raina Deborah, Glory Karimi Kiriinya, Lekonte Salome Mary, Lelikoo Naanyu Sarah, John Kalasinga Lerosion, Abel Senaika, Andrew Solomon, Iliano Loldepe, Longanyek Liliko, Jonathan Letipo, Ruth Kilimo. From World Vision Kenya: Beatrice Mwai, Kevin Muche. Lastly, we would like to thank Jorge Beteta for his translation services.

## SUPPORTING INFORMATION

S1 Tools

S1-S3 Tables, S1-S4 Figs S1

Checklist

