## Supplementary material for "Seasonality, source type, and women’s water labor: A longitudinal mixed-methods study in Kenya and Honduras": S1-S3 Tables, S1-S4 Figs

### **S1 Table.** Demographic and water characteristics of all women who participated in data collection, by country and year

|  | **Kenya** | | | | | | **Honduras** | | | | | | | | | |
| --- | --- | --- | --- | --- | --- | --- | --- | --- | --- | --- | --- | --- | --- | --- | --- | --- |
|  | **2023** | | **2024** | | **Total** | | **2023** | | **2024** | | | | **Total** | | | |
| **Location of water collection/work** | Off premises | | Off premises | | Off premises | | Off premises | | Off premises | | On premises | | Off premises | | Off & On premises | |
| **Total communities** | 6 | | 6 | | 10 | | 6 | | 4 | | 6 | | 8 | | 9 | |
| **Total water collection/work observations** | n = 22 | | n = 48 | | n = 70 | | n = 17 | | n = 11 | | n = 40 | | n = 28 | | n = 68 | |
| **Total unique participants** | n = 22 | | n = 44 | | n = 66 | | n = 17 | | n = 10 | | n = 40 | | n = 27 | | n = 67 | |
| **Total repeat participants** |  | | n = 4 | |  | |  | | n = 1 | |  | | n = 1 | |  | |
| **Age** mean (SD) | 30.2 | (10.1) | 37.1 | (11.7) | 34.9 | (11.6) | 38.9 | (14.5) | 42.4 | (14.3) | 44.7 | (14.3) | 40.3 | (14.3) | 42.9 | (14.3) |
| **Household size** mean (SD) | 6 | (2.3) | 6.1 | (1.8) | 6.1 | (2.0) | 4.2 | (2.2) | 5.3 | (1.6) | 4.0 | (1.6) | 4.6 | (2.1) | 4.3 | (1.8) |
| **Marital status** n, % |  |  |  |  |  |  |  |  |  |  |  |  |  |  |  |  |
| Single, never married | 1 | 4.55 | 1 | 2.08 | 2 | 2.86 | 4 | 23.53 | 1 | 9.09 | 3 | 7.50 | 5 | 17.86 | 8 | 11.76 |
| Unmarried, but have partner | 1 | 4.55 | 0 | 0.00 | 1 | 1.43 | 7 | 41.18 | 6 | 54.55 | 21 | 52.50 | 13 | 46.43 | 34 | 50.00 |
| Married | 17 | 77.27 | 39 | 81.25 | 56 | 80.00 | 3 | 17.65 | 4 | 36.36 | 12 | 30.00 | 7 | 25.00 | 19 | 27.94 |
| Separated | 1 | 4.55 | 0 | 0.00 | 1 | 1.43 | 1 | 5.88 | 0 | 0.00 | 1 | 2.50 | 1 | 3.57 | 2 | 2.94 |
| Widowed | 2 | 9.09 | 8 | 16.67 | 10 | 14.29 | 2 | 11.76 | 0 | 0.00 | 3 | 7.50 | 2 | 7.14 | 5 | 7.35 |
| **Completed schooling^1^** n, % | | | | | | | | | | | | | | | | |
| Never attended school | 20 | 90.91 | 43 | 89.58 | 63 | 90.00 | 4 | 25.00 | 2 | 18.18 | 6 | 15.00 | 6 | 22.22 | 12 | 17.91 |
| Some primary | 0 | 0.00 | 2 | 4.17 | 2 | 2.86 | 6 | 37.50 | 6 | 54.55 | 15 | 37.50 | 12 | 44.44 | 27 | 40.30 |
| Primary | 1 | 4.55 | 2 | 4.17 | 3 | 4.29 | 5 | 31.25 | 3 | 27.27 | 10 | 25.00 | 8 | 29.63 | 18 | 26.87 |
| Some secondary | 0 | 0.00 | 1 | 2.08 | 1 | 1.43 | 0 | 0.00 | 0 | 0.00 | 6 | 15.00 | 0 | 0.00 | 6 | 8.96 |
| Secondary | 0 | 0.00 | 0 | 0.00 | 0 | 0.00 | 1 | 6.25 | 0 | 0.00 | 3 | 7.50 | 1 | 3.70 | 4 | 5.97 |
| Above secondary | 1 | 4.55 | 0 | 0.00 | 1 | 1.43 | 0 | 0.00 | 0 | 0.00 | 0 | 0.00 | 0 | 0.00 | 0 | 0.00 |
| **Engaged in economic activities^1^** n, % | | | | | | | | | | | | | | | | |
| Yes | 6 | 27.27 | 9 | 18.75 | 15 | 21.43 | 6 | 35.29 | 2 | 18.18 | 15 | 38.46 | 8 | 28.57 | 23 | 34.33 |
| No | 16 | 72.73 | 39 | 81.25 | 55 | 78.57 | 11 | 64.71 | 9 | 81.82 | 24 | 61.54 | 20 | 71.43 | 44 | 65.67 |
| **Primary source of household drinking water^2^** n, % | | | | | | | | | | | | | | | | |
| ***Improved*** | ***5*** | ***23.81*** | ***2*** | ***4.17*** | ***7*** | ***10.14*** | ***11*** | ***64.71*** | ***8*** | ***72.73*** | ***34*** | ***87.18*** | ***19*** | ***67.86*** | ***53*** | ***79.10*** |
| Piped water | 0 | 0.00 | 2 | 4.17 | 2 | 2.90 | 7 | 41.18 | 4 | 36.36 | 21 | 53.85 | 11 | 39.29 | 32 | 47.76 |
| Tube well/borehole | 3 | 14.29 | 0 | 0.00 | 3 | 4.35 | 0 | 0.00 | 0 | 0.00 | 0 | 0.00 | 0 | 0.00 | 0 | 0.00 |
| Protected dug well | 2 | 9.52 | 0 | 0.00 | 2 | 2.90 | 1 | 5.88 | 0 | 0.00 | 1 | 2.56 | 1 | 3.57 | 2 | 2.99 |
| Protected spring | 0 | 0.00 | 0 | 0.00 | 0 | 0.00 | 0 | 0.00 | 2 | 18.18 | 2 | 5.13 | 2 | 7.14 | 4 | 5.97 |
| Packaged bottle water | 0 | 0.00 | 0 | 0.00 | 0 | 0.00 | 3 | 17.65 | 0 | 0.00 | 10 | 25.64 | 3 | 10.71 | 13 | 19.40 |
| Packaged sachet water | 0 | 0.00 | 0 | 0.00 | 0 | 0.00 | 0 | 0.00 | 2 | 18.18 | 0 | 0.00 | 2 | 7.14 | 2 | 2.99 |
| ***Unimproved*** | ***16*** | ***76.19*** | ***46*** | ***95.83*** | ***62*** | ***89.86*** | ***6*** | ***35.29*** | ***3*** | ***27.27*** | ***4*** | ***10.26*** | ***9*** | ***32.14*** | ***13*** | ***19.40*** |
| Unprotected dug well | 15 | 71.43 | 26 | 54.17 | 41 | 59.42 | 2 | 11.76 | 1 | 9.09 | 0 | 0.00 | 3 | 10.71 | 3 | 4.48 |
| Unprotected spring | 1 | 4.76 | 0 | 0.00 | 1 | 1.45 | 0 | 0.00 | 2 | 18.18 | 4 | 10.26 | 2 | 7.14 | 6 | 8.96 |
| Surface water | 0 | 0.00 | 20 | 41.67 | 20 | 28.99 | 4 | 23.53 | 0 | 0.00 | 0 | 0.00 | 4 | 14.29 | 4 | 5.97 |
| Other (unspecified) | 0 | 0.00 | 0 | 0.00 | 0 | 0.00 | 0 | 0.00 | 0 | 0.00 | 1 | 2.56 | 0 | 0.00 | 1 | 1.49 |
| **Participant estimated time for household members to collect water, minutes^2^** | | | | | | | | | | | | | | | | |
| mean (SD) | 281.9 | (178.8) | 121.5 | (55.7) | 170.3 | (130.7) | 49.1 | (41.8) | 23.9 | (45.0) | 17.8 | (34.9) | 38.8 | (44.1) | 26.3 | (39.9) |
| **Number of days in week for household members to collect water** | | | | | | | | | | | | | | | | |
| mean (SD) | 5.7 | (1.9) | 5.9 | (1.6) | 5.8 | (1.7) | 4.1 | (2.5) | 4.9 | (2.5) | 4 | (2.6) | 4.4 | (2.5) | 4.2 | (2.5) |
| (min, max) | (2, 7) | | (1, 7) | | (1, 7) | | (1, 7) | | (1, 7) | | (1, 7) | | (1, 7) | | (1, 7) | |
| **Participant estimated total number of trips to water source outside the home on day of data collection** | | | | | | | | | | | | | | | | |
| mean (SD) |  |  | 1.4 | (0.6) |  |  |  |  | 1.1 | (1.1) |  |  |  |  |  |  |
| (min, max) |  |  | (1, 3) | |  |  |  |  | (0, 3) | |  |  |  |  |  |  |
| **Participant estimated water labor time for day of data collection (measured time for those who did water work outside the home water x estimated total number of trips)** | | | | | | | | | | | | | | | | |
| mean (SD) |  |  | 111.3 | (48.8) |  |  |  |  | 49.7 | (47.8) |  |  |  |  |  |  |
| **Participant estimated time to manage water at home for day of data collection** | | | | | | | | | | | | | | | | |
| mean (SD) |  |  | 40.1 | (38.9) |  |  |  |  | 23.3 | (35.7) |  |  |  |  |  |  |
| **Location of drinking water sources^3^** n, % | | | | | | | | | | | | | | | | |
| In dwelling | 0 | 0.00 | 0 | 0.00 | 0 | 0.00 | 1 | 5.88 | 0 | 0.00 | 10 | 25.00 | 1 | 3.70 | 11 | 16.42 |
| In yard/plot | 0 | 0.00 | 0 | 0.00 | 0 | 0.00 | 2 | 11.76 | 2 | 20.00 | 19 | 47.50 | 4 | 14.81 | 23 | 34.33 |
| Elsewhere (beyond yard/plot) | 21 | 100.00 | 47 | 100.00 | 68 | 100.00 | 14 | 82.35 | 8 | 80.00 | 11 | 27.50 | 22 | 81.48 | 33 | 49.25 |
| **Primary source of water for other uses^4^** n, % | | | | | | | | | | | | | | | | |
| ***Improved*** | ***5*** | ***23.81*** | ***4*** | ***8.33*** | ***9*** | ***13.04*** | ***12*** | ***70.59*** | ***6*** | ***54.55*** | ***32*** | ***80.00*** | ***18*** | ***64.29*** | ***50*** | ***73.53*** |
| Piped water | 0 | 0.00 | 2 | 4.17 | 2 | 2.90 | 6 | 35.29 | 3 | 27.27 | 31 | 77.50 | 9 | 32.14 | 40 | 58.82 |
| Tube well/borehole | 3 | 14.29 | 1 | 2.08 | 4 | 5.80 | 0 | 0.00 | 0 | 0.00 | 0 | 0.00 | 0 | 0.00 | 0 | 0.00 |
| Protected dug well | 2 | 9.52 | 0 | 0.00 | 2 | 2.90 | 1 | 5.88 | 1 | 9.09 | 0 | 0.00 | 2 | 7.14 | 2 | 2.94 |
| Protected spring | 0 | 0.00 | 0 | 0.00 | 0 | 0.00 | 0 | 0.00 | 2 | 18.18 | 0 | 0.00 | 2 | 7.14 | 2 | 2.94 |
| Rainwater | 0 | 0.00 | 0 | 0.00 | 0 | 0.00 | 5 | 29.41 | 0 | 0.00 | 1 | 2.50 | 5 | 17.86 | 6 | 8.82 |
| Water kiosk | 0 | 0.00 | 1 | 2.08 | 1 | 1.45 | 0 | 0.00 | 0 | 0.00 | 0 | 0.00 | 0 | 0.00 | 0 | 0.00 |
| ***Unimproved*** | ***16*** | ***76.19*** | ***44*** | ***91.67*** | ***60*** | ***86.96*** | ***5*** | ***29.41*** | ***4*** | ***36.36*** | ***5*** | ***12.50*** | ***9*** | ***32.14*** | ***14*** | ***20.59*** |
| Unprotected dug well | 15 | 71.43 | 25 | 52.08 | 40 | 57.97 | 2 | 11.76 | 0 | 0.00 | 0 | 0.00 | 2 | 7.14 | 2 | 2.94 |
| Unprotected spring | 1 | 4.76 | 0 | 0.00 | 1 | 1.45 | 0 | 0.00 | 0 | 0.00 | 5 | 12.50 | 0 | 0.00 | 5 | 7.35 |
| Surface water | 0 | 0.00 | 19 | 39.58 | 19 | 27.54 | 3 | 17.65 | 4 | 36.36 | 0 | 0.00 | 7 | 25.00 | 7 | 10.29 |
| Other (unspecified) | 0 | 0.00 | 0 | 0.00 | 0 | 0.00 | 0 | 0.00 | 1 | 9.09 | 2 | 5.00 | 1 | 3.57 | 3 | 4.41 |
| No other source | 0 | 0.00 | 0 | 0.00 | 0 | 0.00 | 0 | 0.00 | 0 | 0.00 | 1 | 2.50 | 1 | 2.50 | 1 | 1.47 |
| 1 Honduras: 1 missing  2 Kenya: 2 missing; Honduras: 1 missing  3 Kenya: 3 missing; Honduras: 1 missing  4 Kenya: 1 missing | | | | | | | | | | | | | | | | |

### **S2 Table.** Demographic, water source, and journey characteristics of Kenya analytic and full samples, 2024

|  | **Kenya 2024** | | | |
| --- | --- | --- | --- | --- |
| **Sample** | Analytic sample | | Full sample | |
| **Total communities** | **3** | | **6** | |
| **Total water collection/work observations** | **n = 24**^1^ | | **n = 48**^1^ | |
| **Total unique participants** | n = 20 | | n = 44 | |
| **Total repeat participants** | n = 4 | | n = 4 | |
| **Age** mean (SD) | 37.5 | (14.7) | 37.1 | (11.7) |
| **Household size** mean (SD) | 6.3 | (2.2) | 6.1 | (1.8) |
| **Marital status** n, % |  |  |  |  |
| Single, never married | 1 | 4.17 | 1 | 2.08 |
| Unmarried, but have partner | 0 | 0.00 | 0 | 0.00 |
| Married | 20 | 83.33 | 39 | 81.25 |
| Separated | 0 | 0.00 | 0 | 0.00 |
| Widowed | 3 | 12.50 | 8 | 16.67 |
| **Completed schooling** n, % |  |  |  |  |
| Never attended school | 20 | 83.33 | 43 | 89.58 |
| Some primary | 1 | 4.17 | 2 | 4.17 |
| Primary | 2 | 8.33 | 2 | 4.17 |
| Some secondary | 1 | 4.17 | 1 | 2.08 |
| Secondary | 0 | 0.00 | 0 | 0.00 |
| Above secondary |  |  | 0 | 0.00 |
| **Engaged in economic activities in the last 30 days** n, % |  |  |  |  |
| Yes | 6 | 25.00 | 9 | 18.75 |
| No | 18 | 75.00 | 39 | 81.25 |
| **Primary source of household drinking water** n, % |  |  |  |  |
| ***Improved*** | ***2*** | ***8.33*** | ***2*** | ***4.17*** |
| Piped water | 2 | 8.33 | 2 | 4.17 |
| Tube well/borehole | 0 | 0.00 | 0 | 0.00 |
| Protected dug well | 0 | 0.00 | 0 | 0.00 |
| Protected spring | 0 | 0.00 | 0 | 0.00 |
| Packaged bottle water | 0 | 0.00 | 0 | 0.00 |
| Packaged sachet water | 0 | 0.00 | 0 | 0.00 |
| ***Unimproved*** | ***22*** | ***91.67*** | ***46*** | ***95.83*** |
| Unprotected dug well | 14 | 58.33 | 26 | 54.17 |
| Unprotected spring | 0 | 0.00 | 0 | 0.00 |
| Surface water | 8 | 33.33 | 20 | 41.67 |
| Other (unspecified) | 0 | 0.00 | 0 | 0.00 |
| **Participant estimated time for household members to collect water** (min) | | | | |
| mean (SD) | 106.3 | (49.2) | 121.5 | (55.7) |
| **Number of days in week for household members to collect water** | | | | |
| mean (SD) | 6.3 | (1.4) | 5.9 | (1.6) |
| (min, max) | (1, 7) | | (1, 7) | |
| **Participant estimated total number of trips to water source outside the home on day of data collection** | | | | |
| mean (SD) | 1.6 | (0.7) | 1.4 | (0.6) |
| (min, max) | (0, 3) | | (1, 3) | |
| **Participant estimated water labor time for day of data collection** (min; measured time for those who did water work outside the home x estimated total number of trips) | | | | |
| mean (SD) | 102.7 | (51.8) | 111.3 | (48.8) |
| **Participant estimated time to manage water at home for day of data collection** (min)**^2^** | | | | |
| mean (SD) | 46.4 | (40.1) | 40.1 | (38.9) |
| **Location of drinking water source^3^** n, % |  |  |  |  |
| In dwelling | 0 | 0.00 | 0 | 0.00 |
| In yard/plot | 0 | 0.00 | 0 | 0.00 |
| Elsewhere (beyond yard/plot) | 24 | 100.00 | 47 | 100.00 |
| **Primary source of water for other uses** n, % |  |  |  |  |
| ***Improved*** | ***2*** | ***8.33*** | ***4*** | ***8.33*** |
| Piped water | 1 | 4.17 | 2 | 4.17 |
| Tube well/borehole | 1 | 4.17 | 1 | 2.08 |
| Protected dug well | 0 | 0.00 | 0 | 0.00 |
| Protected spring | 0 | 0.00 | 0 | 0.00 |
| Rainwater | 0 | 0.00 | 0 | 0.00 |
| Water kiosk | 0 | 0.00 | 1 | 2.08 |
| ***Unimproved*** | ***22*** | ***91.67*** | ***44*** | ***91.67*** |
| Unprotected dug well | 14 | 58.33 | 25 | 52.08 |
| Unprotected spring | 0 | 0.00 | 0 | 0.00 |
| Surface water | 8 | 33.33 | 19 | 39.58 |
| Other (unspecified) | 0 | 0.00 | 0 | 0.00 |
| No other source | 0 | 0.00 | 0 | 0.00 |
| **Total time** (min) |  |  |  |  |
| mean (SD) | 74.9 | (41.4) | 90.3 | (40.3) |
| min, max | (19.2 - 150.1) | | (19.2 - 173.4) | |
| **Total distance** (km) |  |  |  |  |
| mean (SD) | 3.4 | (1.8) | 4.1 | (1.7) |
| min, max | (0.5, 5.8) | | (0.5, 6.2) | |
| **Total calories** (kcal) |  |  |  |  |
| mean, SD | 220.3 | (116.7) | 257.8 | (115.2) |
| min, max | (37.0, 405.0) | | (37.0, 507.0) | |
| **Volume of water** (L)**^4^** |  |  |  |  |
| mean (SD) | 22.8 | (7.3) | 22.3 | (6.5) |
| min, max | (10.2, 34.1) | | (10.2, 34.8) | |
| **Volume of water per household member** (L/person)**^4^** |  |  |  |  |
| mean (SD) | 4.1 | (2.0) | 3.9 | (1.7) |
| min, max | (1.6, 10.9) | | (1.6, 10.9) | |
| **Water source type visited during water journey** n, % |  |  |  |  |
| ***Improved*** |  |  |  |  |
| Piped water | 5 | 20.83 | 5 | 10.42 |
| Tube well/borehole | 2 | 8.33 | 2 | 4.17 |
| Water kiosk | 0 | 0.00 | 8 | 16.67 |
| ***Unimproved*** |  |  |  |  |
| Unprotected dug well | 9 | 37.5 | 13 | 27.08 |
| Surface water | 8 | 33.33 | 20 | 41.67 |
| ^1^ 4 women were repeat participants in 2024 ^2^ Kenya analytic sample: 6 missing; Kenya full sample: 10 missing ^3^ Kenya full sample: 1 missing ^4^ Kenya analytic sample: 2 missing; Kenya full sample: 7 missing | | | | |

### **S3 Table.** Journey characteristics of repeat participants in Kenya and Honduras

| **Participant ID** | **Community** | **Year** | **Water source** | **Water source type** | **Activities conducted** | **Total time** (min) | **Total distance** (km) | **Total caloric expenditure** (kcal) | **Other items carried** | **Volume of water**  (1kg water = 1L) | **Volume of water/ person in household** (L/ person) |
| --- | --- | --- | --- | --- | --- | --- | --- | --- | --- | --- | --- |
| **Kenya** |  |  | |  |  |  |  |  |  |  |  |
| K1C | K1 | 2023 | Unimproved | Unprotected dug well | Collected water | 81.9 | 4.9 | 296 | . | 9.1 | 1.1 |
|  | K1 | 2024 | Improved | Piped water (public tank) | Collected water; bathed; collected firewood | 52.9 | 3.2 | 191 | Infant; firewood | 21.5 | 3.1 |
| K1D | K1 | 2023 | Unimproved | Unprotected dug well | Collected water | 106.9 | 6.1 | 302 | . | 24.2 | 3.5 |
|  | K1 | 2024 | Unimproved | Unprotected dug well | Collected water; bathed | 52.0 | 1.9 | 161 | . | 11.0 | 1.6 |
| K1H | K1 | 2023 | Unimproved | Unprotected dug well | Collected water | 117.3 | 5.7 | 294 | . | 3.7 | 0.6 |
|  | K1 | 2024 | Improved | Piped water (public tank) | Collected water | 81.6 | 5.0 | 254 | . | 22.4 | 4.5 |
| K3B | K3 | 2023 | Unimproved | Unprotected dug well | Collected water | 251.4 | 15.8 | 644 | . | . | . |
|  | K3 | 2024 | Unimproved | Surface water (dam) | Collected water | 30.6 | 1.5 | 81 | Infant | 21.7 | 10.9 |
| **Mean 2023** |  |  | |  |  | **139.4** | **8.1** | **384.0** |  | **12.3** | **1.7** |
| **Mean 2024** |  |  | |  |  | **54.3** | **2.9** | **171.8** |  | **19.2** | **5.0** |
| **Mean improved 2024** |  |  | |  |  | **67.2** | **4.1** | **222.5** |  | **22.0** | **3.8** |
| **Mean unimproved 2024** |  |  | |  |  | **41.3** | **1.7** | **121.0** |  | **16.4** | **6.2** |
| **Honduras** |  |  | |  |  |  |  |  |  |  |  |
| Participant H1H | H1 | 2023 | Unimproved | Surface water (river) | Collected water | 30.8 | 1.5 | 106 | . | 17.2 | 2.9 |
|  | H1 | 2024 | Unimproved | Surface water (river) | Washed clothes; bathed child | 101.8 | 1.8 | 221 | Clothes | . | . |

**S1 Fig.** Maps of round-trip routes and activities conducted in 2023 and 2024 among Kenya repeat participants

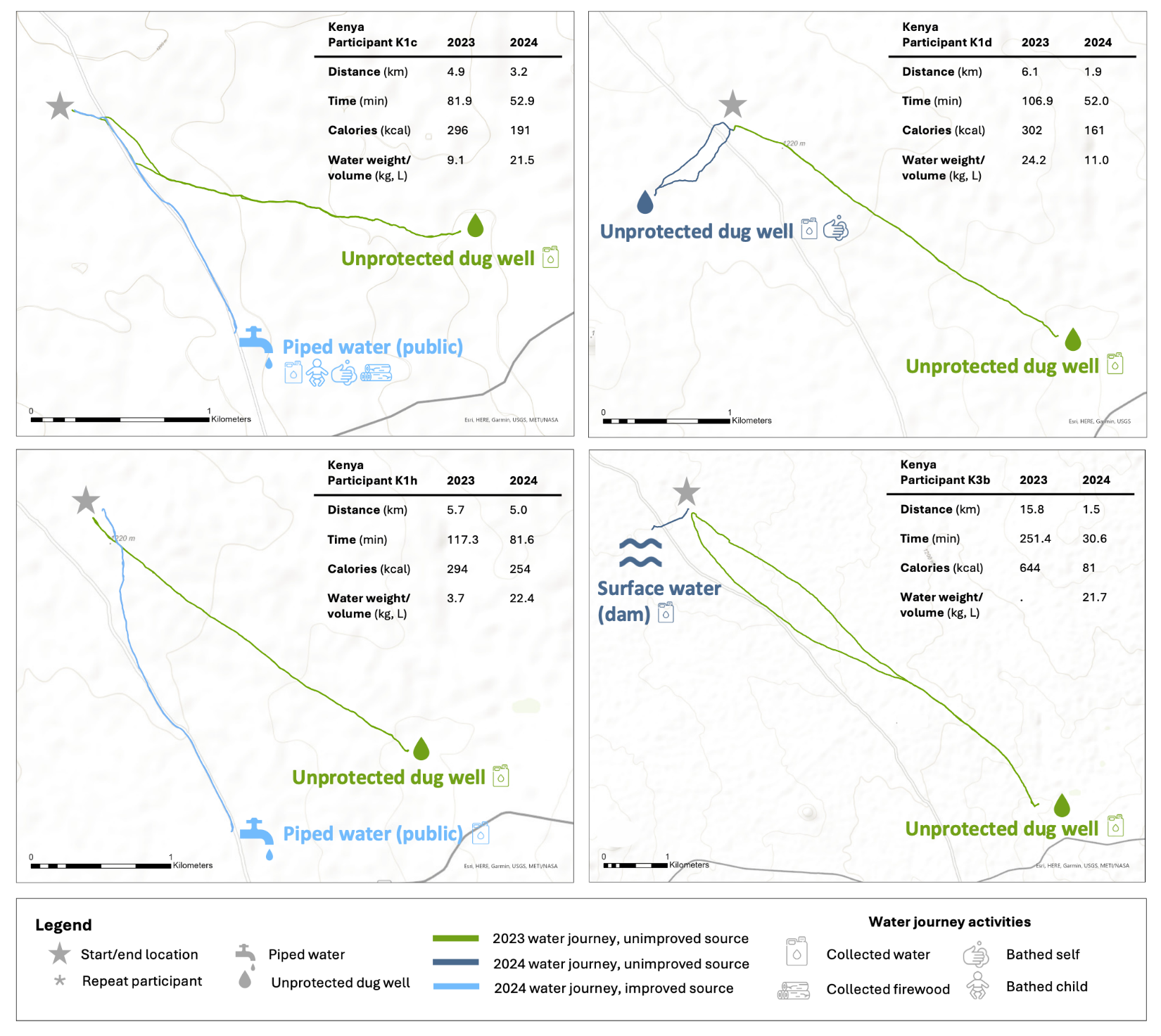

### **S2 Fig.** Map of round-trip routes and activities conducted in 2023 and 2024 for Honduras repeat participant

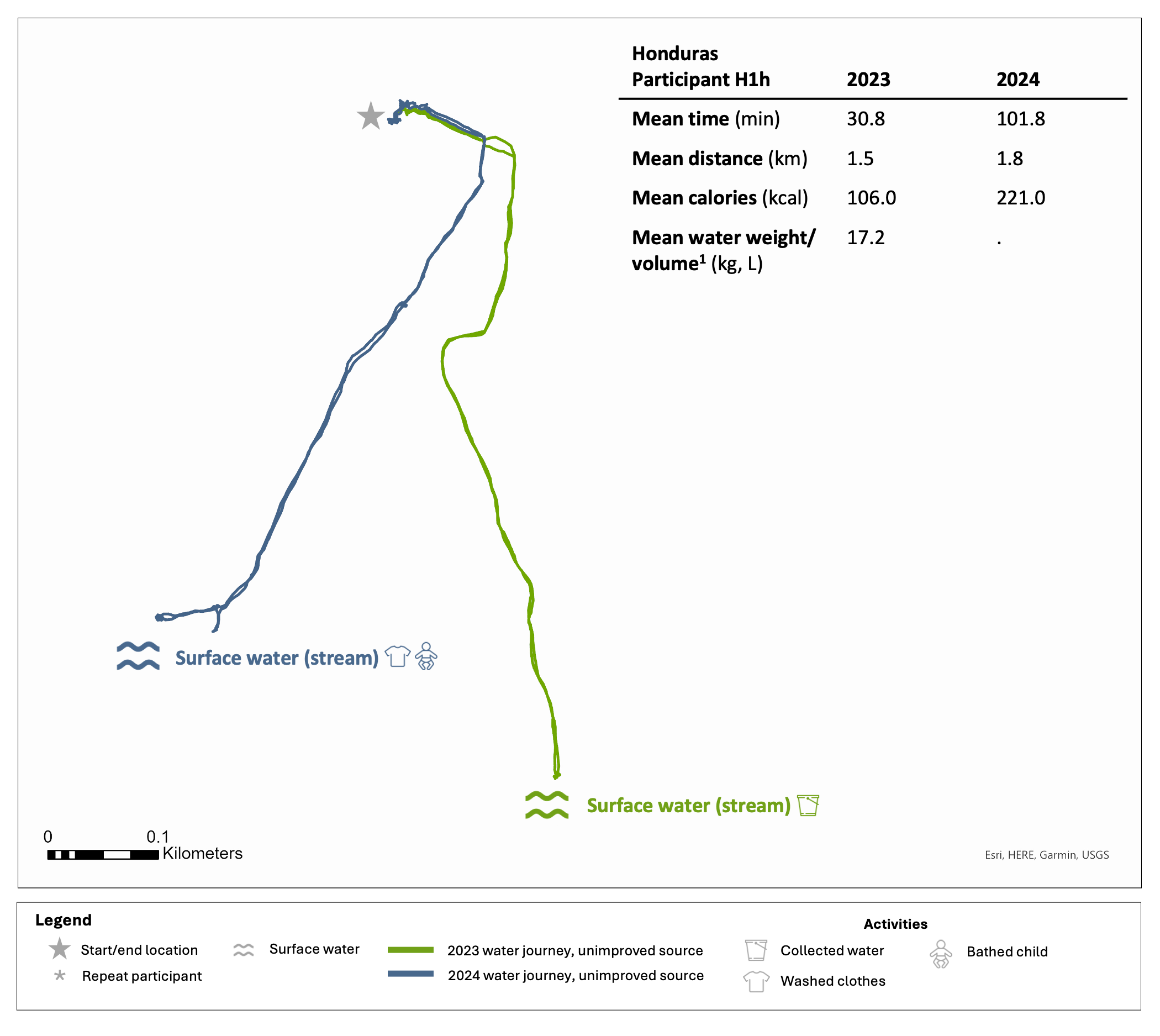

### **S3 Fig.** Time (min) of trips to water source and back among repeat participants in Kenya and Honduras, 2023 and 2024

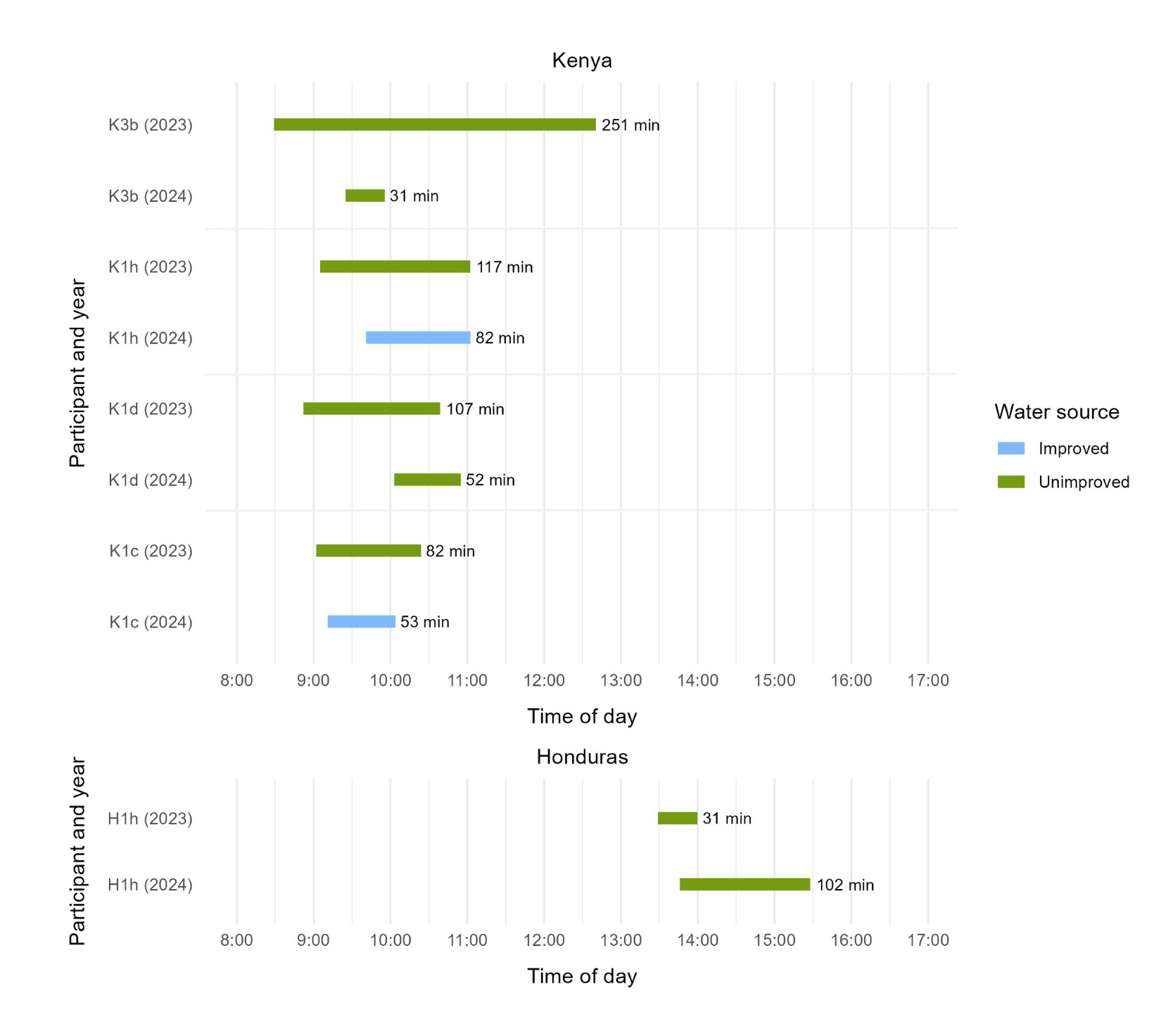

### **S4 Fig.** Mean time (min) of trips to water source and back in Kenya and Honduras, by community and year

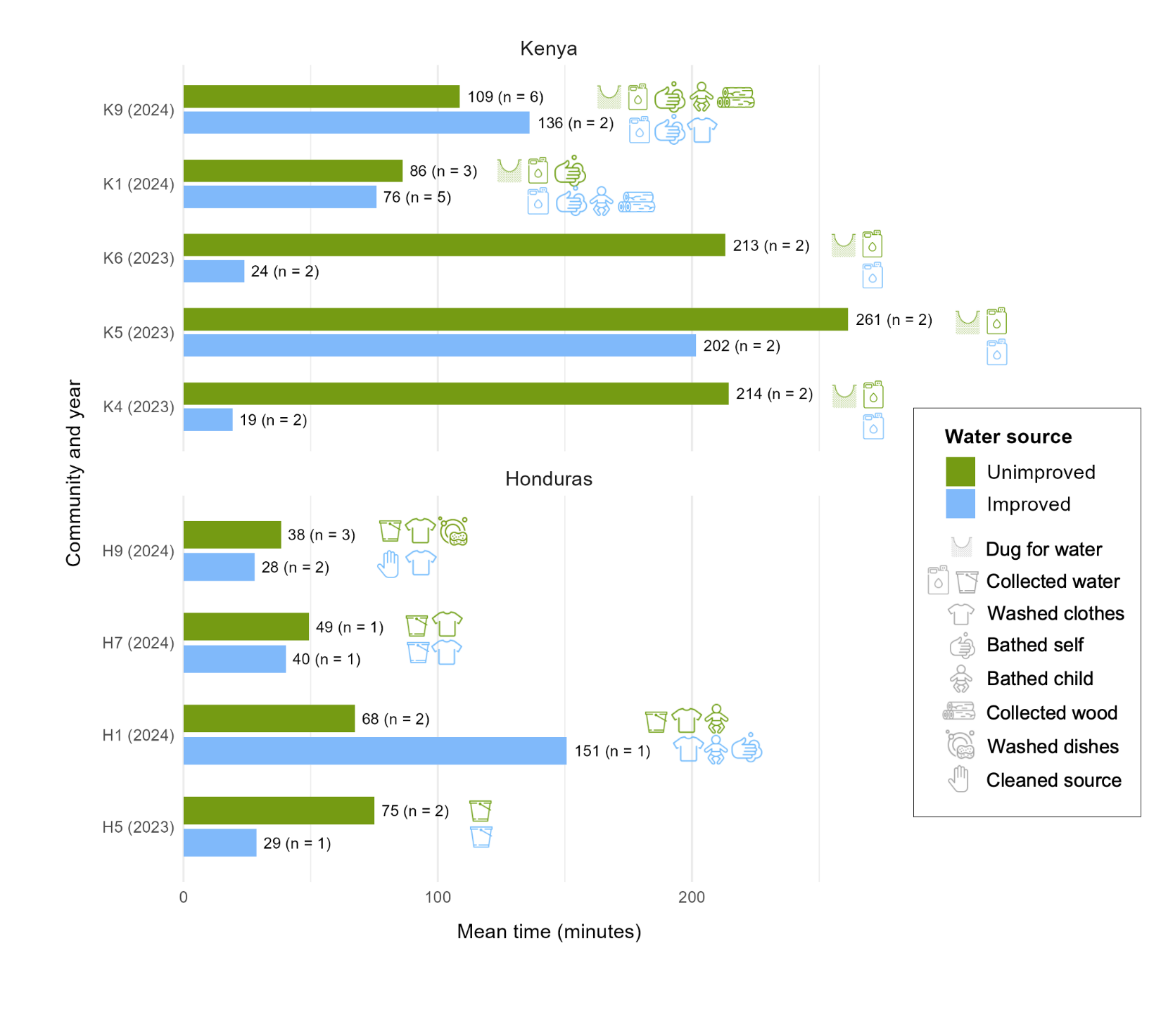
