## Supplementary material for "Seasonality, source type, and women’s water labor: A longitudinal mixed-methods study in Kenya and Honduras": S1 Tools

### **S1a Tools.** Participant Demographic Information, 2023

| **A. Activity information** | | | |
| --- | --- | --- | --- |
| **A001** | Participant ID#: | **A005** | Date: (y/d/m) __ __ __ __ / *__ __ / __ __* |
| **A010** | Community Name: | **A015** | Community ID#: |
| **A017** | **Select activity person will participate in:** | ☐ 1. Key Informant Interview  ☐ 2. Water Journey Go-Along Interview  ☐ 3. FGD with Women  ☐ 4. FGD with Men | |
| **A18** | **If there is more than one activity of this type today, indicate activity number:** | ☐ 1. Not applicable; only activity of this kind here today  ☐ 2. Activity number: **__ __** | |
| **A020** | **Activity Start time:** __ __ : __ __ pm / am | **A025** | **Activity End time:** __ __ : __ __ pm / am |
| **A030** | **Person Filling form**: | **A040** | **Consent Obtained:** ☐ 1. Yes ☐ 2. No |

| **B. Participant Demographic Information** | | | |
| --- | --- | --- | --- |
| **D01** | **Participant Gender**  ☐ 1. Woman ☐ 2. Man | **D02** | **Participant Age __ __**  *(If younger than 18, participant ineligible. End.)* |
| **D03a** | **What is your marital status?**  ☐ 1. Single, never been married  ☐ 2. Unmarried, but have partner  ☐ 3. Married  ☐ 4. Separated  ☐ 5. Divorced  ☐ 6. Widowed | **D03b** | **What type of family structure do you have? [Kenya]**  ☐ 1. Monogamy  ☐ 2. Polygamy |
| **D04** | **What is the highest level of school you completed?**  ☐ 1. Never attended school  ☐ 2. Completed some Primary  ☐ 3. Completed Primary  ☐ 4. Completed Secondary  ☐ 5. Completed schooling above Secondary | | |
| **D05** | **How many people (including yourself) live in your household? __ __**  [Usually number of people sharing meals] | **D06** | **How many children live in your household?**  **__ __**  [This is the total number under age 18.] |
| **D07a** | **Some people take up jobs for which**  **they are paid in cash or kind. Others sell things, have a small business or work on the family farm or in the family business. In the last 30 days, have you done any of these things or any other work?** ☐ 1. Yes ☐ 2. No | **D07b** | ***If participant has engaged in work in the past 30 days (D07a=yes):***  **What are the activities you have done for work?** (write answer) |
| **D08a** | ***If married,* In the last 30 days, has your spouse [husband/wife] done any of these things or any other work?**  ☐ 1. Yes ☐ 2. No | **D08b** | ***If participant’s spouse has engaged in work in the past 30 days (D08a=yes):***  **What are the activities your spouse has done for work?** (write answer) |
| **D09a**  **D09b** | **What is the primary source of DRINKING water used by members of the household in the past 7 days / week?**  **What is the primary source of water FOR OTHER NEEDS used by members of the household in the past 7 days / week?**  **(includes for cooking, bathing, washing clothing, etc.)** | **D09a D09b**  **Drinking Other Uses**  ☐ ☐ 01. No other source used  ☐ ☐ 10. Piped Water  ☐ ☐ 21. Tube well/Borehole  ☐ ☐ 31. Protected Dug Well  ☐ ☐ 32. Unprotected Dug Well  ☐ ☐ 41. Protected Spring  ☐ ☐ 42. Unprotected Spring  ☐ ☐ 51. Rainwater  ☐ ☐ 61. Tanker Truck  ☐ ☐ 71. Cart with small tank  ☐ ☐ 72. Water Kiosk  ☐ ☐ 81. Surface Water *  ☐ ☐ 91. Packaged bottled water  ☐ ☐ 92. Packaged Sachet water  ☐ ☐ 96. Other_____________ *River, dam, lake, pond, stream, canal, irrigation channel | |
| **D10a** | **Where is the household’s main drinking water source?**  ☐ 1. Source in dwelling  ☐ 2. Source in yard/plot  ☐ 3. Source elsewhere (beyond yard/plot) | **D10b** | **How long does it take for members of your household to go to the drinking water source, get water, and come back?**  ☐ 1. __ __ __ minutes  ☐ 88. Do not know  ☐ 99. Not applicable; do not collect drinking water /source on property |
| **D10c** | **How many days in a week does your household collect water from the primary drinking water source?**  __ __  Enter 1-7 for number of days per week  Enter 99 if not applicable  Enter 88 if do not know | **D10d** | **Who is *primarily responsible* for collecting drinking water for the household?**  ☐ 1. Respondent  ☐ 2. Adult woman (age 18 or over)  ☐ 3. Girl (under age 18)  ☐ 4. Adult man (age 18 or over)  ☐ 5. Boy (under age 18)  ☐ 99. Not applicable; do not collect drinking water |
| **D11** | **In the last month, has there been any time when your household did not have sufficient quantities of drinking water when needed?**  ☐ 1. Yes, at least once  ☐ 2. No, Always sufficient  ☐ 88. Do not know | **D11a** | ***If the source for other uses is different than the drinking water source,***  **Where is the household’s main water source for *other uses*?**  ☐ 1. Source in dwelling  ☐ 2. Source in yard/plot  ☐ 3. Source elsewhere (beyond yard/plot) |
| **D11b** | ***If the source for other uses is different than the drinking water source,***  **How long does it take for members of your household to go to the water source for other uses, get water, and come back?**  ☐ 1. __ __ __ minutes  ☐ 88. Do not know  ☐ 99. Not applicable; do not collect drinking water /source on property | **D12** | **Who is *primarily responsible* for collecting water for other uses for the household?**  ☐ 1. Respondent  ☐ 2. Adult woman (age 18 or over)  ☐ 3. Girl (under age 18)  ☐ 4. Adult man (age 18 or over)  ☐ 5. Boy (under age 18)  ☐ 99. Not applicable; do not collect water for other uses |
| **D13a** | **Does your household have access to a toilet facility?**  ☐ 1. Yes, in dwelling  ☐ 2. Yes, in yard/plot  ☐ 3. Yes, elsewhere (beyond yard/plot)  ☐ 4. No household access to a toilet facility | **D13b** | **If your household has access to a toilet facility, do you share it**?  ☐ 1. No, not shared with other households  ☐ 2. Yes, shared with specific households  ☐ 3. Yes, shared with public/community  ☐ 99. Not applicable; no access to a toilet facility |

| **C. Water Journey Information**  *Only collect from Water Journey Participants.* | | | |
| --- | --- | --- | --- |
| **WD01** | **Height:** _ _ _ cm | **WD02** | **Weight: _ _** kg |
| **WD03** | **Date of birth** (yyyy/dd/mm): _ _ _ _ / _ _ / _ _ | | |
| **WD10** | **Length of time lived in community:**  **_ _** years | **WD11** | **Do you intend to leave the community in the next year?**  ☐ 1. Yes  ☐ 2. No  ☐ 88. Do not know |

### **S1b Tools.** Water Journey Go-along Interview Guide, 2023

A. **Background information**

| **Guide: Go-along Interviews and Observations** | | | |
| --- | --- | --- | --- |
| **A010.** | Community Name: | **A015.** | Community ID#: |
| **A020.** | Activity Start time: __ __ : __ __ pm / am | **A025.** | Activity End time: __ __ : __ __ pm / am |
| **A055.** | Date: (y/d/m) __ __ __ __ / *__ __ / __ _* | **A040.** | Consent Obtained: ☐ 1. Yes ☐ 2. No |
| **A045.** | Recorder ID: | **A046.** | Recording #: |
| **A030.** | Interviewer: | **A035.** | Note Taker/Observer: |

B. **Quantitative metrics**

Take the weights of any kind of load that women carry while transporting water. This should include weights of laundry when women do their laundry at the water source.

| **Items** | **Weight (kg)** |
| --- | --- |
| Water containers without water |  |
| Water container with water |  |
| Child(ren) (if carried by participant) |  |
| Laundry (dry) |  |
| Laundry (wet) |  |
| Other (specify): |  |
| Other (specify): |  |
| Other (specify): |  |

C. **Interview questions**

| **Pre and post water collection**   1. Can you walk me through your typical day in relation to water collection? | **Probe:**  What preparations do you make before going to collect water?  **How long does it typically take you to collect water?** |
| --- | --- |
| 2. Where are you currently walking to collect water? | **Probe:**  -What do you plan to use this water for? |
| 3. How often do you collect water for household use at this location (times/week)? |  |
| 4. Where else do you collect water from? | **Probe:**  -Drinking water  -Water for other uses  **Probe**: Laundry, bathing, other  -If water is collected from multiple sources: How do you decide where to collect water from for different activities?  **Probe**: Distance, time, weight, water quality |
| 5. Do you collect water for animal use as well? |  |
| 6. How many trips do you make in a day for water collection? | **Probe:**  -Does this change with seasonality?  -Does this change from day-to-day based on household use (e.g. laundry and other chores that may not be done on a daily basis). |
| 7. What time(s) of the day do you normally go to collect water? | **Probe:**  -Why that time?  -Is there a specific time of the day that you cannot go to the water source? Why?  -If water is collected from multiple sources: repeat question based on source |
| 8. Do you go to the water source by yourself or in a group? Why? | **Probe:**  Do children also go to collect water from these sources? If not, why? |
| 9. Other than water collection, what other activities are you involved in while going to collect water, either along the way or at the water source? | **Probe** (Do not ask probes as yes/no questions; allow participants to describe experiences):  -Socializing with other women  -Fetching firewood on the way  -Picking fruits or vegetables  -Bathing (themselves / children) at the water source  -Washing dishes / laundry at the water source |
| 10. What activities are you involved in to clean or store the water once it has been collected? | **Probe** (Do not ask probes as yes/no questions; allow participants to describe experiences):  -Boiling water  -Filtering water  -Storing water for future use |
| 11. If a water source was brought nearby, how would that change how you manage your time? | **Probe:**  -Productive activities (e.g., garden / agriculture)  -Income-generating activities (e.g., start a business)  **-A**bility to rest |
| 12. What are some of the challenges you experience while collecting water? | **Probe** (Do not ask probes as yes/no questions; allow participants to describe experiences):  -Water scarcity  -Attacks by wild animals  -Attacks by people  -Falls due to terrain  -Having to left heavy containers  -Pain  -Finding childcare or watching children while collecting  -Long lines  -Difficulty walking long distances |
| 13. Do you leave children or other dependents at home while you collect water? | **Probe:**   - Number of children/dependents - Frequency - Do they have supervision? |

### **S1c Tools.** Water Journey Semi-structured Observation Guide, 2023

**Time tracking**

For each activity, document what time the activity begins and ends. Include when the water journey begins, any rest taken along the way, when they arrive at a water point, how long the labor of collection takes, if there is any rest at the water point, when they leave the water point, and when they arrive back at their house.

Some activities, like taking a break, may occur multiple times on the water journey. Document each time it happens. If there are other activities not listed here, make sure you make a note of those as well.

Example:

| **Start Time** | **End Time** | **Activity** |
| --- | --- | --- |
| 8:05 AM | 9:50 AM | Leaves and walks to water collection point |
| 10:50 AM | 11:05 AM | Rests and talks to other women at water point |
| 11:05 AM | 11:45 AM | Pumps water |
| 11:45 AM | 12:00 PM | Rests, feeds child |
| 12:00 PM | 1:30 PM | Walks home with water |
| 1:35 PM | 1:40 PM | Rests, takes drink |
| 1:40 PM | 2:20 PM | Walks home with water |

| **Start Time** | **End Time** | **Activity** |
| --- | --- | --- |

**Qualitative observation**

**Note to observer:** Please document your observations of the water collection journey. We have suggested specific things to pay attention to below, but they are not exhaustive nor are they in order, so please do not limit your observations to what is below. Take notes on the entire journey as you go on it; these notes can be in shorthand as after the observation, you will write out additional notes you did not have time for and debrief with the team. Feel free to also draw maps and pictures as relevant.

| **Observation** | **Detailed description** |
| --- | --- |
| Type and description of containers women use for carrying water | Describe containers (i.e., jerry cans, buckets)  Number of containers  Volume  How the water containers are carried (on head, by handle)  Any other means of transporting water e.g., use of animals like donkeys |
| Health condition and additional items that the woman carry/ bring along to the water source | For example:  Pregnancy  Any visible disability  Carrying a child (i.e., on her back)  Travelling with small animals |
| Terrain | For example:  Hilly, flat, muddy, dry, etc. |
| Water collection process | Describe water access point(s)  Describe water access behavior  When does water collection start, and when does it end?  Who else is at the water source?  Are there are specific areas within the water source specifically meant for women (only women collect water from that point) |
| Any other activity that the woman is engaged in while collecting water | For example:  Taking a break/ rest  Eating  Drinking  Feeding child(ren)  How long are activities? |
| Other observations. Any encounter with wild animals, injuries, encounters with other people |  |

### **S1d Tools.** Participant demographic information survey, 2024

| **A. Activity information** | | | |
| --- | --- | --- | --- |
| **A001** | Participant ID#: | | |
| **A005** | Date: (yyyy/dd/mm) __ __ __ __ / *__ __ / __ __* | | |
| **A010** | Community Name: | | |
| **A015** | Community ID#: | | |
| **A016** | Household ID#: | | |
| **A011** | **Was community engaged for Water Journey data collection in 2023?:**  ☐ 1. Yes ☐ 2. No  *If ‘2.No’ skip to A017.* | **A012** | **If yes, did the participant engage in Water Journey data collection in 2023?**  ☐ 1. Yes ☐ 2. No |
| **A013** | **If yes, what is the participant's**  **Water Journeys 2023 ID#?**  Enumerator note: This information will be provided in advance | 2023 ID#: ________________________ | |
| **A017** | **Select activity person will participate in:** | ☐ 1. Water Journey & Time-Use (Women)  ☐ 2. Water Journey (Women)  ☐ 3. Time-Use (Women)  ☐ 4. Time-Use (Men) | |
| **A020** | **Activity Start time:** __ __ : __ __ pm / am | **A025** | **Activity End time:** __ __ : __ __ pm / am |
| **A030** | Person Filling form: | **A040** | **Confirm Consent Obtained:** ☐ 1. Yes ☐ 2.No  *If* ***no*** *consent was obtained,* ***end*** *participant data collection now.* |

| **B. Participant Demographic Information** | | | |
| --- | --- | --- | --- |
| **D01** | **Participant Gender**  ☐ 1. Woman ☐ 2. Man | **D02** | **What is participant‘s age in complete years?** __ __  *If younger than 18, participant is ineligible.* ***End***  *participant data collection now.* |
| **D03a** | **What is your marital status?**  ☐ 1. Single, never been married  ☐ 2. Unmarried, but have partner  ☐ 3. Married  ☐ 4. Separated  ☐ 5. Divorced  ☐ 6. Widowed | | |
| **D03b** | **(Kenya only)**  **What type of family structure do you have?**  ☐ 1. Monogamy  ☐ 2. Polygamy  ☐ 99. Not applicable | **D03c** | **If married or have a partner, does your spouse/partner live with you?**  ☐ 1. Yes  ☐ 2. No  ☐ 99. Not applicable |
|  |  | **D03d** | **If spouse/partner does not live with you, where do they live?**  ☐ 1. Somewhere else in the same city/town  ☐ 2. Not in this city/town, but in this country  ☐ 3. Not in this country; specify country: ____________________________________  ☐ 99. Not applicable |
| **D04** | **What is the highest level of school you completed?**  ☐ 1. Never attended school  ☐ 2. Completed some Primary  ☐ 3. Completed Primary  ☐ 4. Completed some Secondary  ☐ 5. Completed Secondary  ☐ 6. Completed schooling above Secondary | | |
| **D05** | **How many people (including yourself) live in your household? __ __**  [A household is usually number of people sharing meals] | **D06** | **How many children under age 18 live in your household?**  **__ __** |
| **D07a** | **Some people take up jobs for which they are paid in cash or in exchange for other goods or services. Others sell things, have a small business, or work on the family farm or in the family business.**  **In the last 30 days, have you done any of these things or any other work?**  ☐ 1. Yes ☐ 2. No | **D07b** | *If participant has engaged in paid work in the past 30 days (D07a=yes):*  **What are the activities you have done for paid work?**  **Write answer: _______________________** |
| **D08a** | ***If married or have partner:***  **In the last 30 days, has your spouse/partner done any of these things or any other paid work?**    ☐ 1. Yes ☐ 2. No | **D08b** | ***If participant’s spouse or partner has engaged in paid work in the past 30 days (D08a=yes):***  **What are the activities your spouse/partner has done for paid work?**  **Write answer: _______________________** |
| **D09a** | **What is the primary source of DRINKING water used by members of the household in the past 7 days / week?**  **Enumerators: select only ONE (1) option** | **D09a**  **Drinking**  ☐ 10. Piped Water  ☐ 21. Tube well/Borehole  ☐ 31. Protected Dug Well  ☐ 32. Unprotected Dug Well  (includes digging in riverbed for water, sand abstraction)  ☐ 41. Protected Spring  ☐ 42. Unprotected Spring  ☐ 51. Rainwater  ☐ 61. Tanker Truck  ☐ 71. Cart with small tank  ☐ 72. Water Kiosk  ☐ 81. Surface Water  (River, dam, lake, pond, stream, canal, irrigation channel)  ☐ 91. Packaged bottled water  ☐ 92. Packaged sachet water  ☐ 96. Other ____________________ | |
| **D09b_1** | **What sources of water are used FOR OTHER NEEDS by members of the household in the past 7 days / week?**  (includes for cooking, bathing, washing clothing, etc.)  **Enumerators: Select ALL that apply**  (multiple choice) | **D09b_1**  **Other Uses**  ☐ 01. No other source used  ☐ 10. Piped Water  ☐ 21. Tube well/Borehole  ☐ 31. Protected Dug Well  ☐ 32. Unprotected Dug Well  (includes digging in riverbed for water, Sand abstraction)  ☐ 41. Protected Spring  ☐ 42. Unprotected Spring  ☐ 51. Rainwater  ☐ 61. Tanker Truck  ☐ 71. Cart with small tank  ☐ 72. Water Kiosk  ☐ 81. Surface Water  (River, dam, lake, pond, stream, canal, irrigation channel)  ☐ 91. Packaged bottled water  ☐ 92. Packaged sachet water  ☐ 96. Other _____________________ | |
| **D09b_2** | **What is the primary source of water for OTHER NEEDS?**  Enter ONE (1) number from D09b_1: ______ | | |
| **D09b_3** | **Is the primary source of water for OTHER NEEDS the SAME AS the primary source for DRINKING?**  ☐ 1. Yes ☐ 2. No | | |
| **D10a** | **Where is the household’s main drinking water source?**  ☐ 1. Source in dwelling  ☐ 2. Source in yard/plot  ☐ 3. Source elsewhere (beyond yard/plot) | **D10b** | **How long does it take for members of your household to go to the drinking water source, get water, and come back?**  ☐ 1. __ __ hour(s) __ __ minutes  ☐ 88. Don’t know  ☐ 99. Not applicable; do not collect drinking water /source on property  *Enumerators: if under 1 hour, write 00 for hours* |
| **D10c** | **How many days in a week does your household collect water from the primary drinking water source?**  ______  Enter 1-7 for number of days per week  ☐ 99. Not applicable  ☐ 88. Don’t know | **D10d** | **Who is primarily responsible for collecting drinking water for the household?**  ***Enumerators: Select ONE (1) option***  ☐ 1. Respondent (self)  ☐ 2. Adult woman, other than respondent (age 18 or over)  ☐ 3. Girl (under age 18)  ☐ 4. Adult man, other than respondent  (age 18 or over)  ☐ 5. Boy (under age 18)  ☐ 99. Not applicable; do not collect drinking  water |
| **D11** | **In the last month, has there been any time when your household did not have sufficient quantities of drinking water when needed?**  ☐ 1. Yes, at least once  ☐ 2. No, always sufficient  ☐ 88. Don't know | **D11a** | **Where is the household’s primary water source for OTHER NEEDS?**  **Enumerators:** *this refers to the primary water source for other uses in D09b_2.*  *Fill ‘99’ if D09b_3 = ‘1. Yes.’ Indicating it is the same.*  ☐ 1. Source in dwelling  ☐ 2. Source in yard/plot  ☐ 3. Source elsewhere (beyond yard/plot)  ☐ 99. Not applicable; main source for drinking water and water for other needs are the same |
| **D11b** | **How long does it take for members of your household to go to the primary water source for other needs, get water, and come back?**  **Enumerators:** *this refers to the primary water source for other needs in D09b_2. If same source as drinking water, fill 77.*  ☐ 1. __ __ hour(s) __ __ minutes  ☐ 77. Same as drinking water.  ☐ 88. Don’t know  ☐ 99. Not applicable; do not collect drinking water / source is on property  *Enumerators: if under 1 hour, write 00 for hours* | **D12** | **Who is *primarily responsible* for collecting water for other needs for the household?**  ***Enumerators: Select ONE (1) option***  ☐ 1. Respondent (self)  ☐ 2. Adult woman, other than respondent  (age 18 or over)  ☐ 3. Girl (under age 18)  ☐ 4. Adult man, other than respondent  (age 18 or over)  ☐ 5. Boy (under age 18)  ☐ 99. Not applicable; do not collect water for other needs/source is on property |
| **D13a** | **Does your household have access to a toilet facility?**  ☐ 1. Yes, in dwelling  ☐ 2. Yes, in yard/plot  ☐ 3. Yes, elsewhere (beyond yard/plot)  ☐ 4. No household access to a toilet facility  *If 4 is selected, skip D13b* | **D13b** | **If your household has access to a toilet facility, do you share it**?  ☐ 1. No, not shared with other households  ☐ 2. Yes, shared with specific households  ☐ 3. Yes, shared with public/community  ☐ 99. Not applicable; no access to a toilet facility |

| **C. Water Journey Information**  *Only collect from Water Journey participants (women only)* | | | |
| --- | --- | --- | --- |
| ***Read aloud****: Thank you for providing the earlier information about your household and water source. Now we will take your height and weight as well as ask for your date of birth (or age). This information will help us use the activity-tracking watches during our walk.* | | | |
| **WD01** | *Use tape measure for height*  **Height:** __ __ __ cm | **WD02** | *Use scale for weight*  **Weight:**  __ __ __ . __ __kg |
| **WD03** | **Date of birth** (yyyy/mm/dd): __ __ __ __ / __ __ / __ __  **If date of birth is not known, enter age:** *__ __* years (from D02)  ☐ 88. Don’t know | | |
| **WD10** | **How long have you lived in this community?:**  __ __ years | **WD11** | **Do you intend to leave the community in the next year?**  ☐ 1. Yes  ☐ 2. No  ☐ 88. Don’t know |

### **S1e Tools.** In-depth interview guide, 2024

| **Background (to be filled out by enumerator)** | | | |
| --- | --- | --- | --- |
| **A001.** | Participant ID: ______________________ [*PID should be the same across all modules.]* | | |
| **A010.** | Community Name: | **A015.** | Community ID#: |
| **A016** | Household ID#: |  |  |
| **A020.** | Activity Start time: __ __ : __ __ pm / am | **A025.** | Activity End time: __ __ : __ __ pm / am |
| **A005.** | Date: (y/d/m):  __ __ __ __ / *__ __* / *__ __* | **A040.** | Consent Obtained: ☐ 1. Yes ☐ 2. No  **Enumerators:** If participant says **NO**, **end** data collection now. |
| **A045.** | Recorder ID: | **A046.** | Recording #: |
| **A030.** | Interviewer: | **A035.** | Note Taker/Observer: |

**Module I. Water Journey Interview Questions**

**I.1 Typical water collection**

*Enumerators: These first questions are about the participant’s* ***typical*** *water collection process.*

***READ:*** ***First I will ask you questions about your typical water collection process and experience.***

**I.1.a Please walk me through your typical day collecting water.**

**Probes**

When usually go

When usually return

How often usually go each day

**I.1.b Please describe all the activities you typically collect water for.**

**Probes**

Drinking

Laundry

Cleaning

Bathing

Animal use

**I.1.c How do you typically decide WHERE to collect water?**

**Probe**

Different needs / activities?

Other sources available?

**I.1.d How do you typically decide WHEN to go to collect water?**

**Probes**

Schedule

Temperature

Social time

Safety

**I.1.e How do you typically decide how many times you collect water each day?**

**Probe**

Needs

Weather

Seasons

Source

**I.1.f Please tell me who you typically go with, or meet with, to collect water.**

**Probe**

Children

Community members

**I.1.g What do your children or other family members you care for do when you collect water?**

**Probe**

Number of children/ family members

Need to arrange care/ supervision?

School available for children?

**I.1.h Please explain if you typically pay for water.**

**Probes**

Vary by source?

What is the cost?

How pay? (cash, credit, trade, app)

Who is paid

**I.1.i Please describe any challenges to your typical water collection.**

**Probes**

Water scarcity

Injury / pain

Stress

Childcare

Long lines

Wild animals

**I.1.j Please describe how your water collection practice changes in different seasons.**

**Probe:**

Different source?

More /less time to get to source?

More/less time to retrieve water?

**I.2 Preparations**

*Enumerators: These questions are about the participant’s* ***preparations for*** *water collection.*

***READ:*** ***Now I would like to know about how you prepare to collect water.***

**I.2.a What typical preparations do you make before going to collect water?**

**Probe**

Cleaning containers

Planning other work

Care for family members

Food preparations

**I.2.b When do you make these preparations?**

**Probe**

Morning of

Night before

**I.2.c How long do these preparations usually take?**

**Enumerator enter:** __ __ hour(s) __ __ minutes

**I.3 Other activities while collecting water**

*Enumerators: These questions are about other activities while collecting water.*

***READ:*** ***Now I would like to know what other activities you may do while collecting water.***

**I.3.a Please describe any other activities you typically do while ON THE WAY TO the water source.**

**Probe**

gardening,

socializing,

firewood collection,

Bartering/ selling/ buying goods,

giving water to animals

**I.3.b Please describe any activities you typically do WHILE AT the water source.**

**Probe**

laundry,

bathing,

gardening,

socializing,

firewood collection,

Bartering/ selling/ buying goods,

giving water to animals

**I.3.c Please describe any activities you typically do while ON THE WAY BACK from the water source.**

**Probe**

gardening,

socializing,

firewood collection,

Bartering/ selling/ buying goods,

giving water to animals

**I.4 Water tasks when returning home**

*Enumerators: These questions are about the participant’s* ***activities after*** *water collection.*

***READ:*** ***Now I will ask you questions about what you do with water after you return from collecting.***

**I.4.a What do you typically do with the water you collected when you return home.**

**Probe**

How stored?

Any Treatment? (e.g., boiling, filtering)

Change with season?

**1.4.b Please tell me how long it usually takes you to do all of these water activities when you return home.**

**Enumerator enter:** __ __ hour(s) __ __ minutes

***I.5 If had a better source***

********ONLY for communities that DO NOT yet have a new source from WV *********

*Enumerators: These questions ask participants perceptions of life if they had a better water source.*

***READ:*** ***I am interested to know what you would think of having a new water source.***

**1.5.a Please describe if and how your life would change if a closer and more reliable water source were available to you.**

**Probe**

Time

Activities

Rest / leisure

Income generation

**I.6. New water source**

********ONLY for communities that DO have a new source from WV *********

*Enumerators: These questions ask participants perceptions of life now that they have a new source.*

***READ:*** ***I am interested to know what you think of having a new water source.***

**I.6.a Please describe if and how your life has changed since World Vision built the new source.**

**Probe**

Access

Distance

Time

Reliability

Cost

Social environment

Quality

Availability

**I.6.b Please describe any benefits of having the new water source.**

**I.6.c Please describe any challenges/barriers you have faced with the new water source.**

**I.6.d Do you pay for water at the new source?** ☐ 1. Yes	☐ 2. No

**I.6.e If you pay, what is the cost for what quantity of water?**

**If yes, Indicate cost__________**

**Per quantity_______________**

**I.7 Today’s water collection**

***Enumerators:*** *The following questions are specific to today’s water collection.*

***READ:*** ***Now I have a few more questions about today’s water collection.***

**I.7.a Please describe the water source you will be going to now for water collection.**

**I.7.b Please describe what you have done to prepare for this water collection.**

*Include any activities you did preparing yesterday and today.*

**Probe**

Cleaning water vessels

Completing tasks in the household

Arranging childcare

Cooking

**I.7.c How much time do you think these preparation activities took?**

**Enumerator enter:** __ __ hour(s) __ __ minutes

**I.7.d What you plan to do at this water source?**

**Probe**

Collect water

Laundry

Bathe self/ children

Give animals water

**I.7.e If collecting water to bring home or somewhere else, what do you plan to use this water for?**

**Probes**

Drinking

Cooking

Cleaning

Personal Hygiene /Bathing for self or family

Gardening

Other

**I.7.f What will you do with the water you collect when you return home.**

**Probes**

Store

Use

Treat

**I.7.g How much time do you think you will spend managing your water when you get back home?**

**Enumerator enter:** __ __ hour(s) __ __ minutes

**I.7.h How long do you think this water collection will take you now, including going to the water source, getting water, and returning?**

**Enumerator enter:** __ __ hour(s) __ __ minutes

**I.7.i Have you already been to this water source today?**

**Enumerator enter: __ __ Times**

**I.7.j How many total times do you plan to go to this water source TODAY?**

*Make sure the participant includes visits already made today.*

**Enumerator enter: __ __ Times**

**I.7.k How many times do you plan to go to THIS water source THIS WEEK?**

**Enumerator enter: __ __ Times**

**I.7.l Do you plan to go to ANOTHER water source today?**

☐ 1. Yes, already went	☐ 2. Yes, will go later	☐ 2. Yes, a3ready went and will go again later	☐ 4. No

***If yes, describe source:***

**I.7.m If yes, what did you do or will you do at this other source?**

**I.7.n If yes, how many total times do you plan to go to this OTHER water source TODAY?**

**Enumerator enter: __ __ Times**

**I.7.o If yes, how many times do you plan to go to this OTHER water source THIS WEEK?**

**Enumerator enter: __ __ Times**

**READ:**

**Thank you. I may ask more questions during the journey and have a few more questions when we return. Is there anything else you would like to share before we go to collect water?**

**I.8. Questions at end of the journey**

***Enumerators:*** *The following questions are to be asked after water collection.*

***READ:*** ***Now I have a few final questions about today’s water collection.***

**I.8.a Please describe any activities that you typically do when collecting water that you did not do on this trip.**

***Why did you not do these activities on this trip?***

**Probe**

Bring animals/ children

Laundry

Bathe

**I.8.b Now that you have collected water, how many times do you plan on returning to this or another water source today?**

**Enumerator enter: 1. __ __ Times to this source**

**2. __ __ Times to another source.**

**3. If other source, describe other source _____________________________**

**I.8.b Before we end, please share anything else you would like to share about your experience with water in this community.**

Enumerator: Do not turn off recorder until participant has left.

**READ: Thank you for your time.**

**MODULE W.** Weights of items carried on water journey

Take the weights of any kind of load that women carry while transporting water. This should include weights of laundry when women do their laundry at the water source.

| **Item / person weighed & direction carried** (***CIRCLE*** code to indicate direction) | | | **Enter weight (kg) (fill only one)** | | **Notes** |
| --- | --- | --- | --- | --- | --- |
| **To water source** | **From water source** | **Person or Items weighed** | **Includes participant** (participant alone or holding item/child) | **Does NOT include participant**  (only item/child ) |  |
| W1a. |  | W1. Participant weight | W1c. |  | W1e. |
| W2a. |  | W2. Container(s) without water | W2c. | W2d. | W2e. |
|  | W3b. | W3. Container(s) with water after filling | W3c. | W3d. | W3e. |
| W4a. | W4b. | W4. Child(ren)  (if carried by participant) | W4c. | W4d. | W4e. |
| W5a. | W5b. | W5. Laundry (dry) | W5c. | W5d. | W5e. |
| W6a. | W6b. | W6. Laundry (wet) | W6c. | W6d. | W6e. |
|  |  | W7. Container with water, after arrival home | W7c. | W7d. | W7e. |
| W8a. | W8b. | W8. Other (specify): | W8c. | W8d. | W8e. |
| W9a. | W9b. | W9. Other (specify): | W9c. | W9d. | W9e. |
| W10a. | W10b. | W10. Other (specify): | W10c. | W10d. | W10e. |
| W11a. | W11b. | W11. Other (specify): | W11c. | W11d. | W11e. |
| W12a. | W12b. | W12. Other (specify): | W12c. | W12d. | W12e. |
| *Enumerator note:*  *Other items include* ***any other*** *items carried. This may include food, firewood, items from garden, tools, etc.* | | | | | |
| *Other notes: (If the participant is periodically putting down the child and picking back up, mark that here. If the participant is actually rolling or using a wheelbarrow to carry water, please note here.* | | | | | |

### **S1f Tools.** Semi-structured observation guide, 2024

**T1. Time Tracking For Specific Activities**

The purpose of this chart is to record the times of specific activities. Provide the times for each of the noted activities. Use the column on the far right to add notes about the activities.

| **T1. Times For Specific Activities** | | |
| --- | --- | --- |
| **Add Time** | **Activity** | **Add Notes** |
| T1.1.a | T1.1. Time starts walking to water collection point/source | T1.1.b |
| T1.2.a | T1.2. Time arrives at water collection point/ source | T1.2.b |
| T1.3.a | T1.3. Time starts water collection  (includes pumping, digging, queuing, etc.) | T1.3.b |
| T1.4.a | T1.4. Time finishes water collection  (includes pumping, digging, queuing, etc.) | T1.4.b |
| T1.5.a | T1.5. Time leaves water collection point/source | T1.5.b |
| T1.6.a | T1.6. Time arrive back home or at final destination | T1.6.b |

**T2. Observation-based Activity and Time Tracking**

The purpose of this chart is to understand all activities that take place during the water journey, and how long each takes.

Write down all participant’s activities, the time it begins, and the time it ends. The table should start with when the journey begins and end when the participant returns. Include all activities.

| **T2. EXAMPLE: Observation-based Activity and Time Tracking** | | |
| --- | --- | --- |
| **Start Time** | **End Time** | **Activity** |
| T2.01.a  8:05 AM | T2.01.b  10:50 AM | T2.01  Starts walk. Carries infant on back, rolls 20L can. Child (8) walks and carries 5L can |
| T2.02.a  10:50 AM | T2.02.b  11:05 AM | T2.02  Waits in line, talks to other women at water point, breastfeeds infant |
| T2.03.a  11:05 AM | T2.03.b  11:45 AM | T2.03  Pumps water |
| T2.04.a  11:45 AM | T2.04.b  12:00 PM | T2.04  Rests, drinks water, gives water to 8-year old, breastfeeds infant |
| T2.05.a  12:00 | T2.05.b  1:30 | T2.05  Starts home, 20 L container on back and infant on front. Child carries 5 L can. |
| T2.06.a  1:30 PM | T2.06.b  1:50 PM | T2.06  Hands infant to 8-year old to carry on back; carries both jerry cans |
| T2.07.a  1:55 PM | T2.07.b  1:57 PM | T2.07  Picks up firewood |
| T2.08.a  12:57 PM | T2.08.b  2:09 PM | T2.08  Rests, takes drink |
| T2.09.a  2:09 PM | T2.09.b  3:12 PM | T2.09  Continues walk with 10L jerry can on back, firewood secured on top. Rolls 20 L can |
| T2.10.a  3:12 PM | T2.10.b  3:12 | T2.10  Arrives back home |

| **T2. Observation-based Activity and Time Tracking** | | |
| --- | --- | --- |
| **Add**  **Start Time** | **Add**  **End Time** | **Describe Activity** |
| T2.01.a | T2.01.b | T2.01:  Leaves home to walk to water collection point. |
| T2.02.a | T2.02.b | T2.02 |
| T2.03.a | T2.03.b | T2.03 |
| T2.04.a | T2.04.b | T2.04 |
| T2.05.a | T2.05.b | T2.05 |
| T2.06.a | T2.06.b | T2.06 |
| T2.07.a | T2.07.b | T2.07 |
| T2.08.a | T2.08.b | T2.08 |
| T2.09.a | T2.09.b | T2.09 |
| T2.10.a | T2.10.b | T2.10 |
| T2.11.a | T2.11.b | T2.11 |
| T2.12.a | T2.12.b | T2.12 |
| T2.13.a | T2.13.b | T2.13 |
| T2.14.a | T2.14.b | T2.14 |
| T2.15.a | T2.15.b | T2.15 |
| T2.16.a | T2.16.b | T2.16 |
| T2.17.a | T2.17.b | T2.17 |
| T2.18.a | T2.18.b | T2.18 |

**Observation Prompts**

**O1. Describe containers woman uses to collect water**

Note each, one by one, including: type (jerry can, buckets, etc.); volume; how carried [on head, rolled behind participant, etc.]; any other means of transport [donkey, wheelbarrow])

*Example:*

1. *Jerry can, 20L, strapped on back*
2. *Jerry can, 10L, carried on head*

**O2. Describe additional items being carried or brought with participant** **to water point**

- May include: children (note walking /being carried); animals (type & number); goods to barter; firewood; laundry; fertilizer; etc.
- Note how that item is being carried, and if carried the entire time (children may walk some).
- *All items noted here should have weight recorded in module W.*

**O3. Provide physical description of participant**

Note: if pregnant; if any disability, illness, or injury; what kind of shoes is she wearing.

**O4. Describe Terrain** **on the way to the water point**

Note: if there a road and/or path to walk on; if uphill, downhill, hilly, flat, muddy, slippery, rocky, dry, etc.

Take notes as the terrain changes, for example, if they start on a flat road but then get walk down a rocky hill.

**O5. Note any activities the participant carried out on the way to water point**

*(Examples: collecting firewood, tending garden, feeding child, resting, buying/selling goods, etc.)*

**O6. Describe the water point, including:**

- **Type** (such as river, dug well, borehole)
- **Terrain** around water source (sandy, muddy, rocky, bottom of hill, etc. )
- **Presence of others** (note if is it busy, if there other women, men, children, animals (wild or livestock) there using the water point, if there separate areas for men and women

**O8.** **Document the process of water collection.** Processes may include:

- Waiting in line; digging; being handed water up from the well; using a mechanical pump; working with other people to pump; paying a water vendor;

**O9. Note any other activities the participant engaged in at water point**

*Activities may include:* eating, drinking, resting, feeding children, socializing, bathing, bathing children, washing clothes, washing utensils, bartering/buying/selling goods, et.c)

**O10. Describe how full water containers are when the participant starts the return jouney, and note the transport method**

Note if containers women use for carrying water are completely or only partially full; if any left behind for later transport. Note how containers are carried back (on head, on back, with wheelbarrow, etc.) and if this method changes at any point.

**O11. Note all non-water items being brought back with participant from water point.**

- May include: children (note walking or being carried); animals (type & number); goods to barter; firewood; laundry; fertilizer; etc.
- Note how that item is being carried, and if carried the entire time (children may walk some).
- All items noted here should have weight recorded in Module W.

**O12. Describe Terrain ON THE WAY FROM the water point**

Note: This could be different than the way there as it is the opposite direction. And the participant may have gone a different way. Note if it is the same or a different way. As with the journey to the source, note if there a road and/or path to walk on; if uphill, downhill, hilly, flat, muddy, slippery, rocky, dry, etc. Take notes as the terrain shifts and changes – for example, if they start on a flat road but then get off to walk down a rocky hill.

**O13. Note any activities the participant carried out on the way to water point**

*(Examples: collecting firewood, tending garden, feeding child, resting, buying/selling goods, etc.)*

**O14. Note the status of woman during and after the journey.**

Does she seem tired, energetic, stressed; does weight seem to be very burdensome/ easy for her to carry? etc.)

Any injuries?

**O15. Any notable encounters with wild animals? other people?**

**O16. Other enumerator notes: Is there anything you observed that we should add to this guide? Any other notes?**
